# Characterization of SARS-CoV-2 vaccine waning in Mexico

**DOI:** 10.1101/2022.04.17.22273854

**Authors:** Carlos Hernandez-Suarez, Efren Murillo-Zamora

**Author notes:** Email addresses:* (Carlos Hernandez-Suarez), (Efren Murillo-Zamora).

## Abstract

We use survival analysis to analyze the decay in the protection induced by eight vaccines using data from 44, 006 patients from the IMSS public health system in Mexico, including only previously vaccinated, confirmed SARS-CoV-2 positive with a PCR test. We analyze three groupings: all data, complete vs. incomplete dose and less than 60 years or older. We found that a Weibull distribution fits very well the complete dose data. Only three vaccines still had 30% of their initial strength after 32 weeks. In two-dose vaccines, we found that the average protection time of a complete dose increases 2 to 3 times compared to that of an incomplete dose.

## 2. Introduction

Several reports suggest that the protective effect of vaccines against SARS-CoV-2 virus wanes with time. Characterizing how immunity wanes is relevant for policy making, especially regarding vaccination strategies (Goldberg et al., 2021). These kind of studies may be useful to design the best interval between doses or booster shots.

Studies that analyze how the vaccine protection decays over time can be classified into two types: those that measure surrogates of humoral response and those that measure the vaccine efficacy (VE) at several successive points in time. Among the first category is Housset et al. (2022) who observed a decrease in anti-spike antibody titer of 84.3% between months 1-6 for Pfizer vaccine, whereas (Levine-Tiefenbrun et al., 2022) analyzed the cycle threshold (Ct) values of RdRp gene, that initially increased by 2.7 relative to unvaccinated in the first month after the booster dose, but then decayed to 1.3 in the second month and found to be small in the third to fourth months. Health care workers –considered to be at higher risk– who received the two doses of the Pfizer vaccine developed protective antibodies that were maintained at detectable levels at least for 250 days after the second dose of the vaccine (Coppeta et al., 2022). Other studies that report reduction in humoral response for several vaccines include Berar-Yanay et al. (2021); Kolaric et al. (2021); Khoury et al. (2021); Peled et al.; Levin et al. (2021)

The estimates of VE are in general *relative risk* measures, some function of attack ratios (Lin et al., 2022) measured usually through a cohort-study or a test-negative design (Fukushima & Hirota, 2017) and thus provide a comparison between relative risk of vaccinated and unvaccinated groups. Tartof et al. (2021) argues that a reduction in vaccine effectiveness against SARS-CoV-2 infections over time is more likely to be due to waning immunity rather than the delta variant escaping vaccine protection. Several studies suggest a significant waning of VE from 90 days after the second dose (Kurita et al., 2022; Andrews et al., 2022; Bedston et al., 2022; Ferdinands et al., 2022; Goldberg et al., 2021).

In here, we use survival analysis in which event times are the time to infection from the application of the first dose. The advantage of using survival analysis is that the decay of the protective effect is not a relative measure, as it occurs with VE but instead is a measure of how the protective effect decays independently of the response of the individual when the protection fails. This will allow to propose an index to measure the protective effect of the vaccine that is not a comparative measure with non-vaccinated.

### Vaccines included in the study

Table 1 shows the vaccines included in this work, whereas Table 2 shows the amount of data available for each vaccine in the first 14 months.

**Table 1:** Vaccine names, manufacturer an abbreviations used in this work.

| Vaccine | Manufacturer | Abbreviation |
| --- | --- | --- |
| AZD1222 | Oxford/AstraZeneca | AZ |
| Ad5-nCoV Convidecia | CanSino | CA |
| mRNA-1273 | Moderna Biotech | MO |
| BBIBP-CorV | Sinopharm | SP |
| CoronaVac | Sinovac | SV |
| Sputnik V/Gam-COVID-Vac | Gamaleya | GA |
| Ad26.COV2. S | Janssen | JA |
| BNT162b2 | Pfizer/BioNTech | PF |

**Table 2:** Number of vaccinated individuals exceeding *n* months from their vaccination with a first dose up to 29/Mar/2022.

| Months | AZ | CA | MO | SP | SV | GA | JA | PF |
| --- | --- | --- | --- | --- | --- | --- | --- | --- |
| 3 | 499,528 | 64,681 | 24,111 | 6,925 | 129,441 | 89,351 | 60,861 | 354,110 |
| 4 | 495,515 | 64,139 | 23,927 | 6,912 | 129,105 | 88,748 | 60,723 | 349,870 |
| 5 | 485,542 | 62,466 | 23,337 | 6,848 | 127,840 | 86,386 | 60,465 | 344,890 |
| 6 | 467,213 | 58,711 | 21,441 | 6,644 | 125,222 | 72,846 | 60,104 | 338,373 |
| 7 | 432,815 | 56,224 | 11,496 | 6,308 | 119,469 | 62,808 | 59,279 | 321,107 |
| 8 | 344,027 | 50,940 | 5,657 | 4,817 | 91,977 | 56,368 | 57,670 | 292,447 |
| 9 | 191,611 | 45,854 | 3,696 | 2,738 | 47,583 | 31,549 | 51,998 | 246,435 |
| 10 | 96,637 | 41,605 | 2,552 | 1,839 | 30,490 | 23,232 | 7,478 | 202,420 |
| 11 | 47,542 | 22,493 | 1,497 | 1,137 | 17,783 | 11,942 | 1,160 | 147,750 |
| 12 | 25,326 | 6,704 | 802 | 812 | 11,923 | 6,466 | 602 | 116,041 |
| 13 | 12,125 | 2,000 | 428 | 311 | 3,519 | 2,872 | 253 | 84,354 |
| 14 | 4,353 | 467 | 204 | 71 | 787 | 638 | 102 | 52,014 |

### The data

The Mexican Institute for Social Insurance (IMSS) had in February 2022 about 8 M insured (IMSS, 2022). The IMSS has a COVID-19 surveillance system (SINOLAVE) that recorded 5’365,955 cases from 29-Dec-2019 to 01-Apr-2022. From this database we extracted vaccinated individuals with a PCR positive test for SARS-CoV-2. We considered only the first confirmed infection in case an individual had two or more confirmed reinfections. Figure 2 shows the data selection process, from the original data set to the working database with 44,006 observations and 17 variables. Some basics statistics of this later database are shown in Table 3, whereas Table 4 shows statistics on the number of vaccinated individuals for each vaccine in our study.

**Table 3:** Age frequencies of Data set 4 (see Figure 2) by gender and outcome.

| Age category |  | Gender |  | Hospitalized |  |  | Home |
| --- | --- | --- | --- | --- | --- | --- | --- |
| L | U | Male | Female | Dead | Recovered | n.d.* |  |
| 0 | 15 | 38 | 73 | - | 12 | 3 | 96 |
| 16 | 30 | 4,065 | 6,331 | 72 | 608 | 70 | 9,646 |
| 31 | 59 | 10,614 | 13,394 | 1,584 | 2,907 | 320 | 19,197 |
| 60 | — | 5,416 | 4,075 | 3,911 | 3,000 | 391 | 2,189 |

\*Non-determined up to 29/Mar/2022.

**Table 4:**
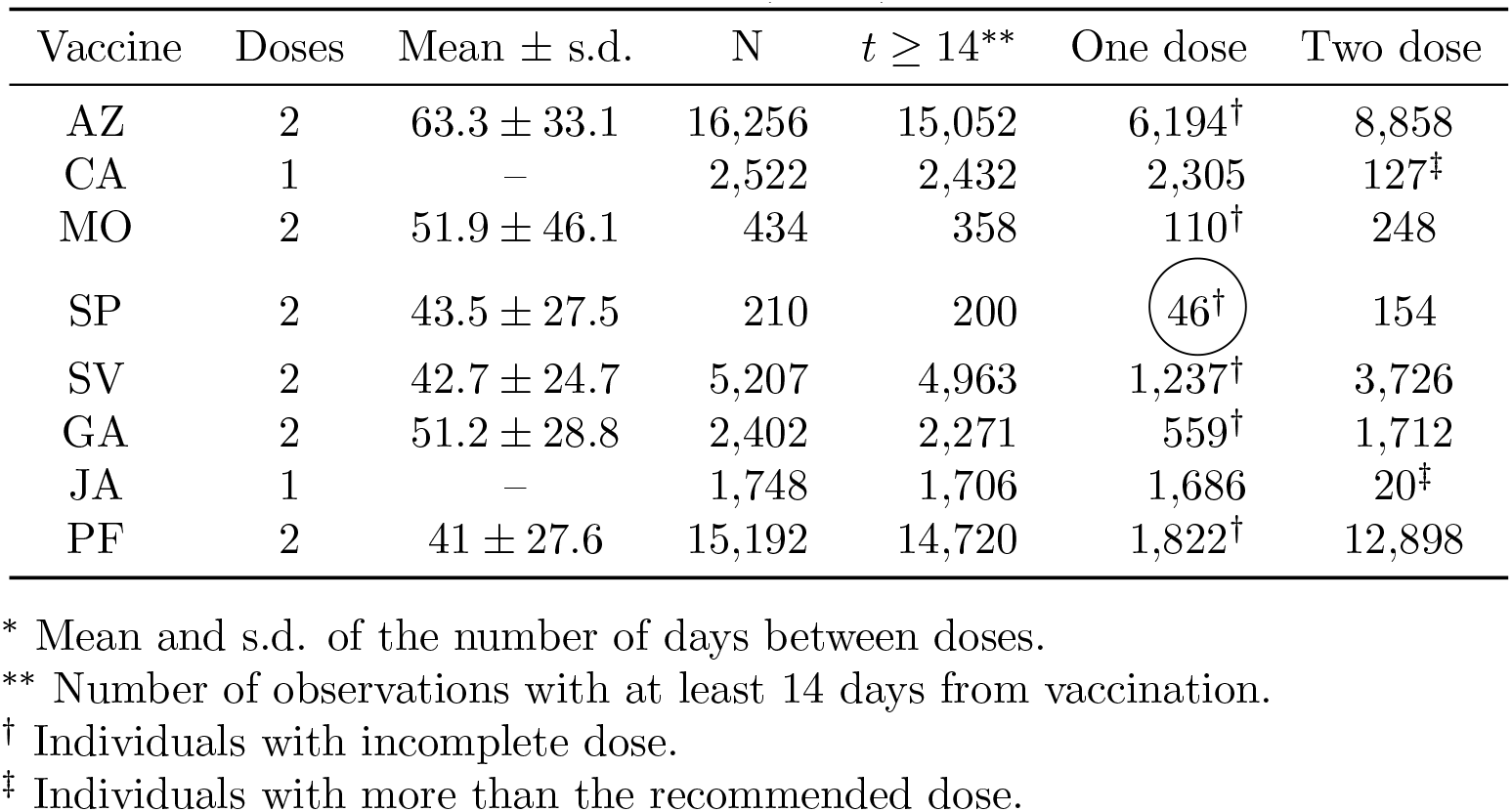
Number of infections observed for every vaccine. A Weibull distributed was not fitted in those cases with less than 100 observations (circled).

| Vaccine | Doses | Mean ± s.d. | N | $t \geq 14^{**}$ | One dose | Two dose |
| --- | --- | --- | --- | --- | --- | --- |
| AZ | 2 | $63.3 \pm 33.1$ | 16,256 | 15,052 | 6,194<sup>†</sup> | 8,858 |
| CA | 1 | – | 2,522 | 2,432 | 2,305 | 127<sup>‡</sup> |
| MO | 2 | $51.9 \pm 46.1$ | 434 | 358 | 110<sup>†</sup> | 248 |
| SP | 2 | $43.5 \pm 27.5$ | 210 | 200 | 46<sup>†</sup> | 154 |
| SV | 2 | $42.7 \pm 24.7$ | 5,207 | 4,963 | 1,237<sup>†</sup> | 3,726 |
| GA | 2 | $51.2 \pm 28.8$ | 2,402 | 2,271 | 559<sup>†</sup> | 1,712 |
| JA | 1 | – | 1,748 | 1,706 | 1,686 | 20<sup>‡</sup> |
| PF | 2 | $41 \pm 27.6$ | 15,192 | 14,720 | 1,822<sup>†</sup> | 12,898 |

\* Mean and s.d. of the number of days between doses.
\*\* Number of observations with at least 14 days from vaccination.
<sup>†</sup> Individuals with incomplete dose.
<sup>‡</sup> Individuals with more than the recommended dose.

### Methodology

The first vaccine was applied on 07/Ene/2021, and the database ends on 29/Mar/2022. For each vaccinated individual that was eventually confirmed to be infected with SARS-CoV-2 with a PCR test, we used the day of vaccination and the day when symptoms started. It is reasonable to assume that if an individual is vaccinated at time *t*_0_, the protective effect of a vaccine *d* days later depends only on *d* and not on *t*_0_, therefore, the data to characterize the waning effect of a particular vaccine *v* has the form *t*_1_, *t*_2_, *t*_3_, …, *N*_*v*_ where *t*_*i*_ is the elapsed time from vaccination to beginning of symptoms and *N*_*v*_ is the total number of individuals confirmed infected vaccinated with vaccine *v*. We eliminated those infections occurring in the first two weeks from vaccination, when apparently not enough immune response has been built (Afshar et al.).

We disregard all individuals whose infection was undetected or that were not infected from their vaccination day to 9/Mar/2022. Censorship does not affect the estimation process other than the reduction in sample size as long as every observation is equally likely to be excluded (Leung et al., 1997). We now discuss the effect of this censorship in the analysis.

### 2.1. Justifying the use of non-censored observations exclusively

Basically, if a fraction *p* of the individuals is eliminated from the analysis, this does not have more implications for the estimation process other than the reduction in sample size, as long as every individual has the same probability *p* of being excluded from the sample. In our case, we have two kinds of individuals eliminated from the analysis: a) those whose infection was not detected and b) those that were not infected between the time from first vaccination until 29/Mar/2022.

Regarding case (a), it is more likely that those individuals with mild responses are less prone to seek for help at a hospital and thus time of infection from vaccination of mild cases may be underrepresented in the sample. If the (unobserved) infections including both mild and severe cases occur at times *t*_1_, *t*_2_, *t*_3_, … and mild cases are distributed at random among these observations, then even if we exclude all mild cases this would not produce a bias in the estimation process (Leung et al., 1997). To evaluate the possibility of non-random allocation of mild/severe cases, we plot the times of infection of all vaccinated individuals with a positive PCR test as a function of time from vaccination, in two groups: hospitalized (severe cases) and ambulatory (mild cases). Figure 1 shows how the cumulative proportion of hospitalized and ambulatory cases evolve with time from vaccination. Although it is impossible to know the fraction of mild cases lost and when they were lost, it seems that at least for the first ten months from vaccination the ratio mild to severe cases detected is preserved, which favors the argument that appearance of mild/sever cases are random and thus, mild cases are lost at random.

**Figure 1:**
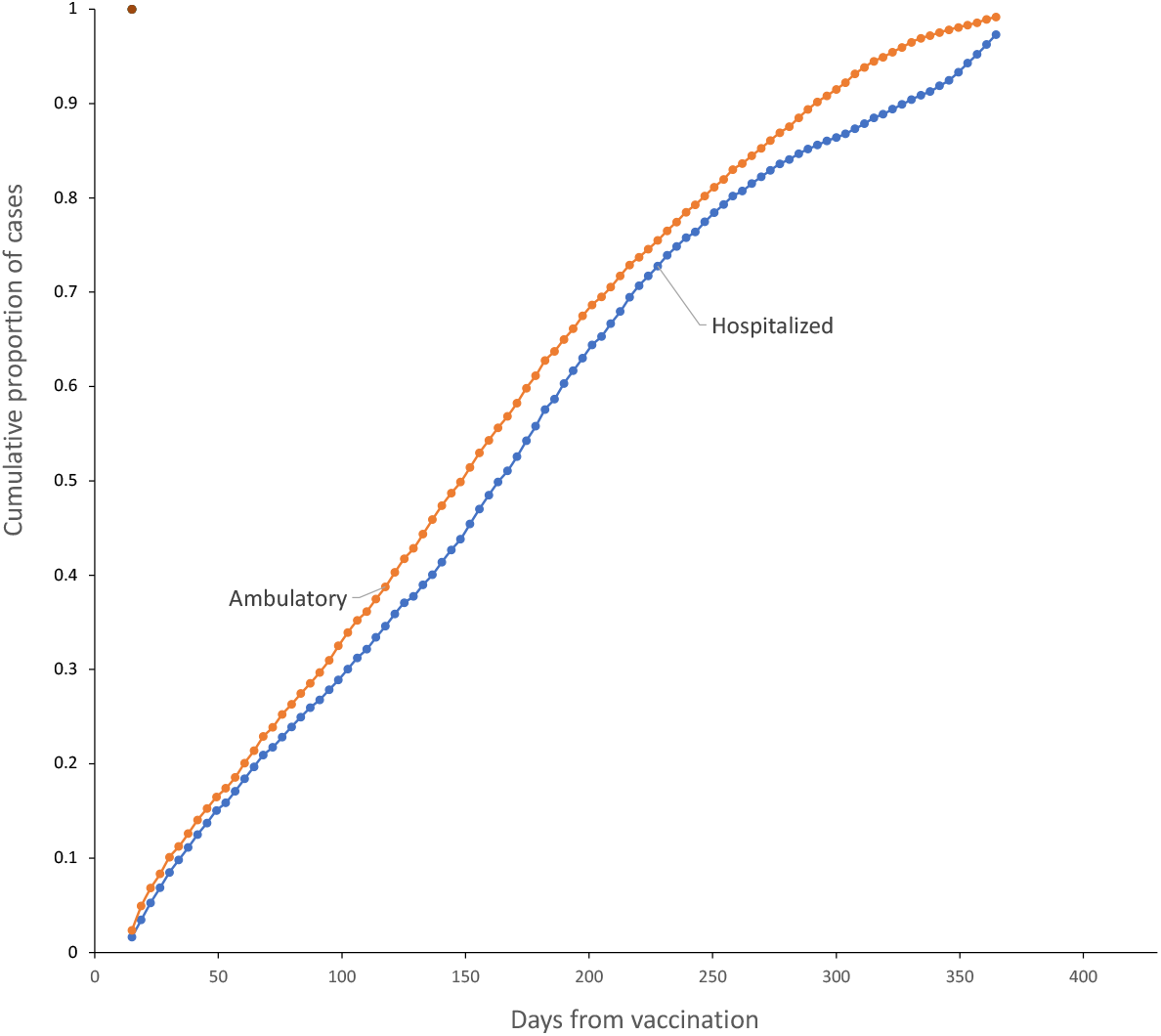
Evolution of the cumulative proportions of hospitalized (red) and ambulatory (blue) among the vaccinated infected, as a function of time from vaccination.

If the case is (b), in which vaccinated individuals were not infected from the day of first shot to 29/Mar/2022, they may reflect a protective effect of a vaccine, and thus, avoiding them adds a bias to the estimate, since an individual would tend to be eliminated from the sample if a vaccine confers a great amount of protection for a long time. There is a way to reduce the bias caused by this kind of censorship: chose a value of *t*^*^ large enough so that there is a high certainty that no vaccine protection exceeds *t*^*^, and censor the observations that were not followed for at least *t*^*^ days, unless the event occurred before *t*^*^. The amount of bias introduced by this method depends on how large is *t*^*^, and the bias is 0 if *t* → ∞ or if we wait until all vaccinated are infected. Since the largest time elapsed between first vaccine and the appearance of symptoms in our data is 434 days, this is the maximum value we can choose for *t*^*^, which is a reasonable amount of time for the vanishing vaccine effect, according to the literature. This approach directly censors all vaccinated with no detected infection because they were not infected before *t*^*^ or did not reached *t*^*^ units of observation.

The method used here is robust to the presence of individuals with natural immunity among the vaccinated. That is, if there are individuals which are immune to the disease among those vaccinated, the waning effect of vaccines can still be parameterized. We assume that the vaccine provides some amount of initial protection against infection and with time this protection wanes and infections will start occurring among those vaccinated. If there are immune individuals among vaccinated, they are still immune when the vaccine effect wanes completely.

Observe that *t*_*i*_ and *t*_*j*_ may be correlated, but they are independent, which is essential for estimation purposes. We cannot record the exact day of infection, and will use the day in which symptoms started, as declared by the patients.

As it will be shown later, the data suggests a failure time according to a Weibull distribution, with *cumulative hazard function* (*λt*)^*k*^. The Weibull distribution is a versatile distribution that has been extensively used in destructive processes in industry and medicine (Johnson et al., 2005). We will obtain maximum likelihood estimates (MLE’s) of *λ* and *k*.

### Measuring vaccine performance

It is important to emphasize that this analysis only allows to characterize how the protective effect wanes with time, regardless of the amount of initial protection conferred by the vaccine. A vaccine may induce a stronger protection than another at the beginning, but its protective effect may wane faster, and, at the end, it could offer less total protective effect. Observe that:

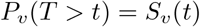

is the probability that an individual receiving vaccine *v* is not infected in the next *t* days after first vaccination. With this, we suggest using as a measure performance the index:

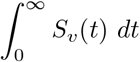

which is the total protection conferred by vaccine *v* during its lifespan. Observe that by definition, the previous integral is the expected value of *T*. Therefore, the average time to infection under vaccine *v* will be used as a measure of performance.

## 3. Results

We adjusted a Weibull distribution to each vaccine, leading to a *survival function*:

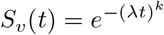

which has a mean *λ* Γ(1+1*/k*) where Γ(*·*) is the *Gamma* mathematical function.

### 3.1. A single group for every vaccine: all observations included

In a first analysis, we include all observations in 5-th column of Table 4, that is, all individuals with infection time of at least 14 days after first shot, with or without complete dose. This will be useful to compare with the disaggregated behavior for complete and incomplete dose. The fitted parameter estimates along the expected time to infection are shown in Table 5.

**Table 5:** Fitted parameters for the survival function *S*(*t*) for a Weibull distribution (*λ, k*) and other statistics for each vaccine analyzed. Data from complete and incomplete dose is considered.

| Vaccine | $\lambda$ | $k$ | $N$ | $R^2$ | $\mu^*$ | Average<sup>†</sup> |
| --- | --- | --- | --- | --- | --- | --- |
| AZ | 147.52 | 1.50 | 15,052 | 1.0 | 133.2 | 124.0 |
| CA | 163.37 | 1.95 | 2,305 | 1.0 | 144.9 | 139.8 |
| MO | 162.37 | 1.79 | 358 | 1.0 | 144.4 | 121.2 |
| SP | 171.16 | 1.98 | 200 | 1.0 | 151.7 | 145.4 |
| SV | 158.15 | 1.74 | 4,963 | 1.0 | 140.9 | 135.2 |
| GA | 155.37 | 1.87 | 2,271 | 1.0 | 138.0 | 131.0 |
| JA | 181.96 | 2.67 | 1,686 | 1.0 | 161.8 | 158.8 |
| PF | 245.07 | 2.15 | 14,720 | 1.0 | 217.0 | 211.9 |

\* $\mu = \lambda \Gamma(1 + k)$ , from adjusted Weibull model.
<sup>†</sup> From data.

The observed survival curves for each vaccine are shown in Figure 3 and the observed survival curve for each vaccine together with the fitted Weibull survival distribution are shown in Figures 4-11.

4in

### 3.2. Disaggregating complete vs. incomplete dose

In this section we analyze observed vs. adjusted survival curves for each vaccine, considering whether the individuals completed the recommended number of doses or not. Table 4 shows the amount of data available for each group (last two columns). We can analyze the effect of a complete dose in all eight vaccines but only five allow to study the effect of an incomplete dose (see Table 4).

In particular for the CA vaccine, there were 127 individuals with two dose (see Table 4), that is, one more than the recommended. Nevertheless, the time elapsed between both doses is very irregular to be considered: the median of the elapsed time for these individuals is 44.5 days, with only 14 individuals receiving the second dose within 4 weeks and 64 within 8 weeks. There were 20 individuals with JA vaccine with more than the single recommended dose, which were also excluded.

Table 6 shows the parameters for the adjusted Weibull model for each case, and the observed survival curve together with the fitted Weibull survival distribution for both complete and incomplete dose are shown in Figures 12-19. Confidence intervals are provided in the Appendix (10).

**Table 6:** Fitted parameters for the survival function *S*(*t*) for a Weibull distribution (*λ, k*) for complete and incomplete dose.

| Vaccine | Complete dose |  |  |  | Incomplete dose |  |  |  |  |
| --- | --- | --- | --- | --- | --- | --- | --- | --- | --- |
| | $N_1$ | $\lambda_1$ | $k_1$ | $\mu_1^*$ | $N_0$ | $\lambda_0$ | $k_0$ | $\mu_0^*$ | $\mu_1/\mu_0$ |
| AZ | 8,858 | 207.1 | 2.8 | 184.4 | 6,194 | 66.5 | 1.4 | 61 | 3 |
| CA | 2,305 | 163.4 | 2 | 144.9 | - | 0 | 0 | - | - |
| MO | 248 | 195.2 | 2.8 | 173.9 | 110 | 85 | 1.1 | 81 | 2.1 |
| SP | 154 | 196.8 | 2.8 | 175.2 | 46 | 0 | 0 | - | - |
| SV | 3,726 | 188.2 | 2.6 | 167.1 | 1,237 | 68.4 | 1.2 | 64.7 | 2.6 |
| GA | 1,712 | 181.1 | 2.5 | 160.7 | 559 | 76.6 | 1.3 | 70.6 | 2.3 |
| JA | 1,686 | 182 | 2.7 | 161.8 | - | 0 | 0 | - | - |
| PF | 12,898 | 266 | 2.8 | 236.9 | 1,822 | 91.8 | 1.1 | 88.9 | 2.7 |

\* $\mu_i = \lambda \Gamma(1 + k)$ , from adjusted Weibull model.

### 3.3. Disaggregating by age

Now, for all individuals with a complete dose, we disaggregate those with 60 years or older against the rest and adjusted a survival curve to each vaccine by group age. The adjusted survival curves are shown in Figures 20-26. Table 7 shows the adjusted parameters in each case. Confidence intervals are provided in the Appendix (10,11).

**Table 7:** Fitted parameters for the survival function *S*(*t*) for a Weibull distribution (*λ, k*) for age *≥* 60 and age < 60. Complete dose only.

| Vaccine | Age $< 60$ | | | | Age $\geq 60$ | | | |
| --- | --- | --- | --- | --- | --- | --- | --- | --- |
| | $N_0$ | $\lambda_0$ | $k_1$ | $\mu_0$ | $N_1$ | $\lambda_1$ | $k_1$ | $\mu_1$ |
| AZ | 6,503 | 201.9 | 2.9 | 180.0 | 2,355 | 220.9 | 2.6 | 196.2 |
| CA* | 1,735 | 161.5 | 1.9 | 143.2 | 570 | 169.1 | 2 | 149.9 |
| MO | 228 | 194.3 | 2.8 | 173.0 | 20 | – | – | – |
| SP | 78 | – | – | – | 76 | – | – | – |
| SV | 2,181 | 176 | 2.6 | 156.3 | 1,545 | 205.1 | 2.7 | 182.4 |
| GA | 1,345 | 173 | 2.6 | 153.6 | 367 | 209.7 | 2.5 | 186.1 |
| JA | 1,635 | 181.7 | 2.7 | 161.5 | 51 | – | – | – |
| PF† | 10,028 | 273.3 | 2.9 | 243.6 | 2,870 | 239.7 | 2.8 | 213.5 |

\* Does not reject $H_0 : \mu_1 = \mu_0$ .
† Does not reject $H_0 : \mu_0 > \mu_1$ .

## 4. Discussion

Different individuals may react differently when the vaccine strength has been reduced, say, to 50%, depending on facts as comorbidities, amount of exposure, age, gender, etc. For some, a 50% reduction level represents still a high level of protection whereas for others it is already a huge loss. The methods used here can be applied to more specific categories as the ones described (age, complete/incomplete dose). The waning effect is summarized in Tables 8 and 9 including only vaccines with at least 300 observations.

**Table 8:** Day at loss of a given protection ability of vaccines among those receiving complete dose with age < 60 years, with at least 300 observations. Calculated from the adjusted Weibull distribution

| Vaccine | N | Proportion of protection lost |  |  |  |  |  |
| --- | --- | --- | --- | --- | --- | --- | --- |
|  |  | 0.5 | 0.6 | 0.7 | 0.8 | 0.9 | 0.95 |
| AZ | 6,503 | 178 | 196 | 215 | 238 | 269 | 295 |
| CA | 1,735 | 133 | 154 | 178 | 207 | 251 | 288 |
| MO | 228 | — | — | — | — | — | — |
| SP | 78 | — | — | — | — | — | — |
| SV | 2,181 | 153 | 170 | 189 | 211 | 243 | 268 |
| GA | 1,345 | 150 | 167 | 186 | 208 | 238 | 264 |
| JA | 1,635 | 159 | 176 | 195 | 217 | 247 | 273 |
| PF | 10,028 | 241 | 265 | 291 | 322 | 364 | 399 |

**Table 9:**
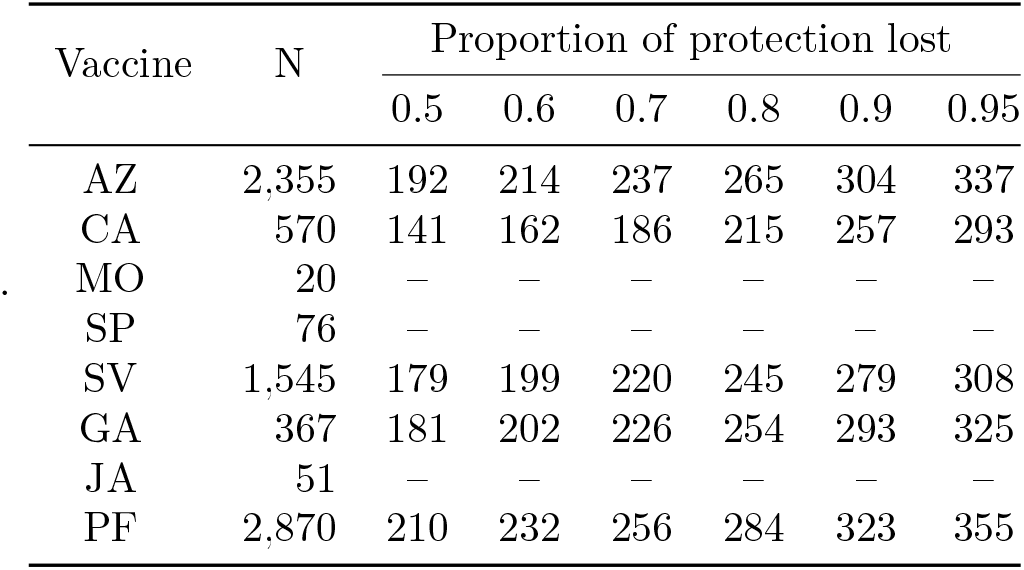
Day at loss of a given protection ability of vaccines among those receiving complete dose with age < 60 years, with at least 300 observations. Calculated from the adjusted Weibull distribution

The main difference with other studies to characterize the waning effect of vaccines is that in this study we consider the time to failure, instead of a sample taken at regular intervals. This allows for a parameterization of the survival curves *S*_*v*_(*t*). The Weibull model fits remarkably well, specially considering those factors mentioned previously, as the potential effect of differential exposure. The results in this work agree with the claims that most of the protective ability has waned after seven months, which is true for most vaccines in the study. Although the fitted model can be used to obtain the average time to infection from vaccination, special care must be taken with predicting in the boundaries. For instance, the individuals that had a complete dose with the PF vaccine had in average 236.7 days to infection from first dose, whereas the adjusted complete dose model (Table 6) shows that *µ* = 236.9, which is quite similar. Nevertheless, 95% of individuals vaccinated with PF vaccine had been infected by day 369, whereas the Weibull model with the parameter for complete dose of Table 6 predicts that 95% of individuals will lost the vaccine protection by day 393, a difference of 24 days. We chose this example because the data exhibit a fast decay of PF vaccine from a year of vaccination (see Fig. 19) which the adjusted Weibull model did not capture.

In general, the models for complete dose seem to have better fit than those for incomplete dose, although there is also the possibility that these later require a different survival function. The usefulness of the incomplete dose data depends on its nature: if the individual misses the second dose for a reason that is associated with greater exposure, as it is the case of individuals that may have lost interest in receiving the second dose or believe that they have enough protection already, those may be factors that increase the exposure (Blower & McLean, 1994) and thus the data on incomplete dose would be useless. If the lack of a second dose is independent from exposure or risk, then the incomplete dose model provides information on the increase achieved by observing the recommendations of the manufacturers. The ratio *µ*_1_*/µ*_0_ in Table 6 suggests that the increase in average protection time of a complete dose is between 2 −3 times that of an incomplete dose. In all cases, the null hypothesis that then mean of both groups is equal is rejected with *p* < 0.001.

When disaggregated by age, we observed that the protection waned faster in general, in younger people. One may be tempted to believe that this differential protection between both age groups may be due to a greater exposure of one of the groups, but if this is the case, the behavior should be consistent across vaccines. What we observed is that the CA vaccine did not reject the hypothesis of equal means and that the PF vaccine the younger group had a larger mean protection than the older. For the rest, the older group had longer protection than the younger (see Tables 7, 8 and 9). This lack of consistency may suggest an interaction vaccine-age group.

The role of differential pressure needs to be analyzed more deeply. Clearly, higher infectious pressure may produce an infection quicker than under low infection pressure, but we do not know the functional relationship between infection pressure and the increase in risk of infection. For this, we analyzed the deployment of every vaccine through time: upon defining the *vaccine age* of an individual as the current time from vaccination with the first dose, then, it is possible to analyze the *vaccine age* through time. These *vaccine ages* can be compared with some surrogate of infection pressure. In this case, we use as a surrogate the confirmed cases for Mexico using data from Dong et al. (2020). For instance, Figure 27 shows that in the middle of August 21, there was a peak in confirmed daily cases of SARS-CoV-2, and by that time the individuals with the PF vaccine had the largest average *vaccine age* (141 d), whereas those vaccinated with the CA vaccine had the lowest average (58 d). This plot can give us an idea on whether a vaccine has been deployed during higher infection pressure or not.

Although we are measuring decay in vaccine protection, this should have a correspondence with VE of the form *V E* ∝ *S*(*t*). Nordstrom et al. (2022) obtained estimates of VE for several cohorts for vaccines PF, MO, AZ and a mixture of AZ and MO. Unfortunately, the intervals are too width to be comparable in some cases. For instance, Nordstrom et al. (2022) reports that AZ vaccine has a VE of 49% in the interval 31-60 day after second doses, and 41% in the interval 61-120 days. This implies that the 45% VE is reached somewhere between 31 - 120 days. For PF vaccine, Nordstrom et al. (2022) reports that an estimated VE of 47% was observed in the interval of 121-180 days after second dose. Using the complete model parameters in Table 6, PF vaccine loses 50% protective effect by day 233 from first dose, subtracting the average time to second dose (Table 4) of 41 days for this vaccine, yields a loss of 50% the protective effect by day 192.

Also, for PF vaccine, Nordstrom et al. (2022) reports a VE of 23% after 210 days from second dose. The Weibull model for the complete dose (Table 6) shows that a loss of 77% protection is achieved by day 305 from first dose or 264 days from second dose. A larger contrast between Nordstrom et al. (2022) an this study is related to the AZ vaccine, since they found this vaccine has lost the VE after 120 days from second dose, while our complete dose model suggests that, after subtracting the average time between first and second dose, by day 118 AZ still exhibits a 50% protective strength.

In more agreement with Nordstrom et al. (2022) are their results when all groups were combined: a VE of 50% was observed by days 121-180 after second dose, and that a VE of 23% was estimated after day 210. In our analysis, when combining all vaccines and weighting for sample size, we found that a 50% protective effect is lost by day 151 from second dose and that 75% efficacy has been lost by day 210.

Our results also agree with those of Andrews et al. (2022), where it is reported that by day 140 after second dose, the vaccines AZ and PF vaccines have reduced the VE to 44.3% and 66.3% respectively. When using the survival curve from our adjusted Weibull model for the complete dose, the protective efficacy of the vaccines by day 140 has been reduced to 38% and 71% respectively. Also Bedston et al. (2022) estimates a VE of 86% by day 14 and of 53% by day 154 for PF vaccine after second dose, whereas our survival model for complete dose PF vaccine for those days is 99% and 66% respectively.

It is important to notice that there were 1, 528 vaccinated individuals with a *vaccine age* larger than 400 days and of these, only 40 (2.6%) gave a positive PCR in that interval and out of these later group, 4 (10%) where hospitalized. There is an interesting interpretation of these facts, since they show that when the vaccine effect had reasonably waned for all vaccines, only less than 0.3% required hospitalization. Given the high infectiousness of the different SARS-CoV-2 variants, this may suggest that there may be a large proportion of individuals with natural immunity in the population.

## Data Availability

All data produced in the present study are available upon reasonable request to the authors

## 5. Ethics declaration

This study was reviewed and approved by the Local Health Research Committee 601 of the Instituto Mexicano de Seguro Social (approval R-2020-601-022).

## 7. Appendix

**Table 10:**
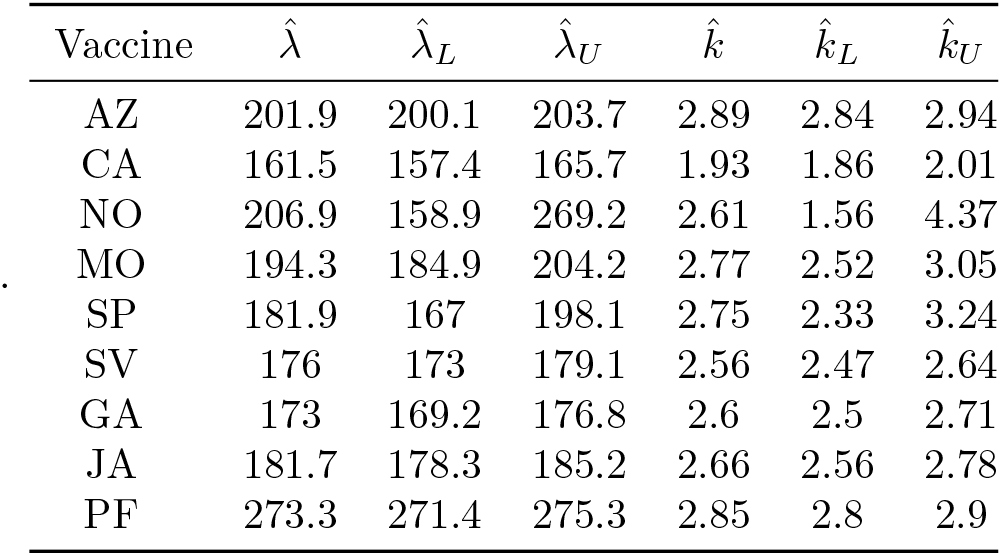
Estimates and 95% CI for the parameters of the Weibull distribution, for complete dose only with age < 60

**Table 11:** Estimates and 95% CI for the parameters of the Weibull distribution, for complete dose only with age ≥ 60

| Vaccine | $\hat{\lambda}$ | $\hat{\lambda}_L$ | $\hat{\lambda}_U$ | $\hat{k}$ | $\hat{k}_L$ | $\hat{k}_U$ |
| --- | --- | --- | --- | --- | --- | --- |
| AZ | 220.9 | 217.3 | 224.5 | 2.62 | 2.53 | 2.7 |
| CA | 169.1 | 162 | 176.5 | 2.01 | 1.88 | 2.14 |
| NO | 228.7 | 187.5 | 278.8 | 2.89 | 1.87 | 4.46 |
| MO | 204.7 | 181.7 | 230.7 | 3.89 | 2.75 | 5.48 |
| SP | 211.9 | 195 | 230.2 | 2.88 | 2.43 | 3.42 |
| SV | 205.1 | 201.1 | 209.1 | 2.7 | 2.6 | 2.8 |
| GA | 209.7 | 200.9 | 218.9 | 2.52 | 2.32 | 2.73 |
| JA | 189 | 170.9 | 208.9 | 2.85 | 2.3 | 3.54 |
| PF | 239.7 | 236.4 | 242.9 | 2.84 | 2.76 | 2.93 |

**Figure 2:**
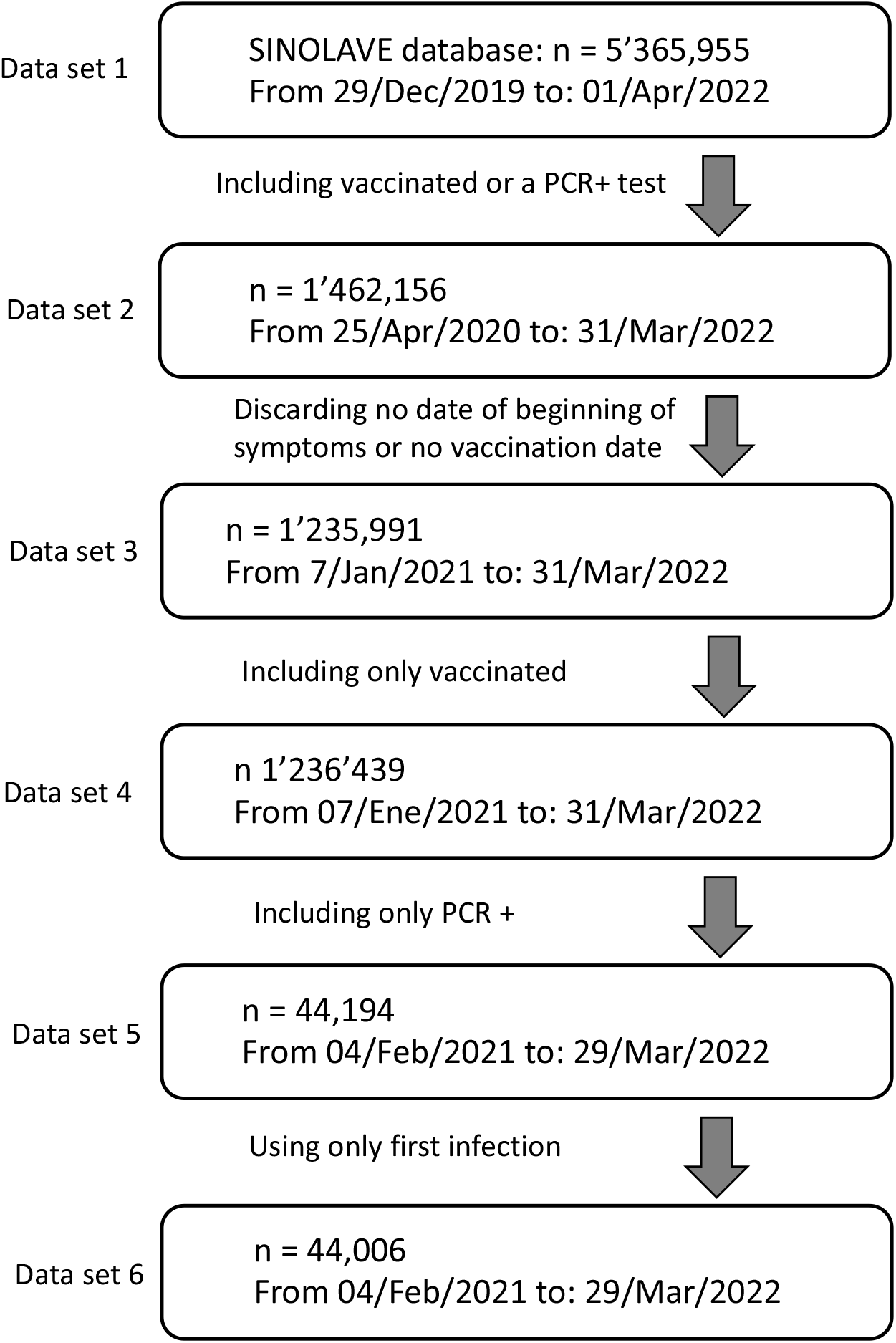
Data selection process from the original SINOLAVE IMSS database with 5’365,955 observations and 169 variables to the working database with 44,006 observations and 17 variables. The data obtained at each step is required for additional analysis not included here.

**Figure 3:**
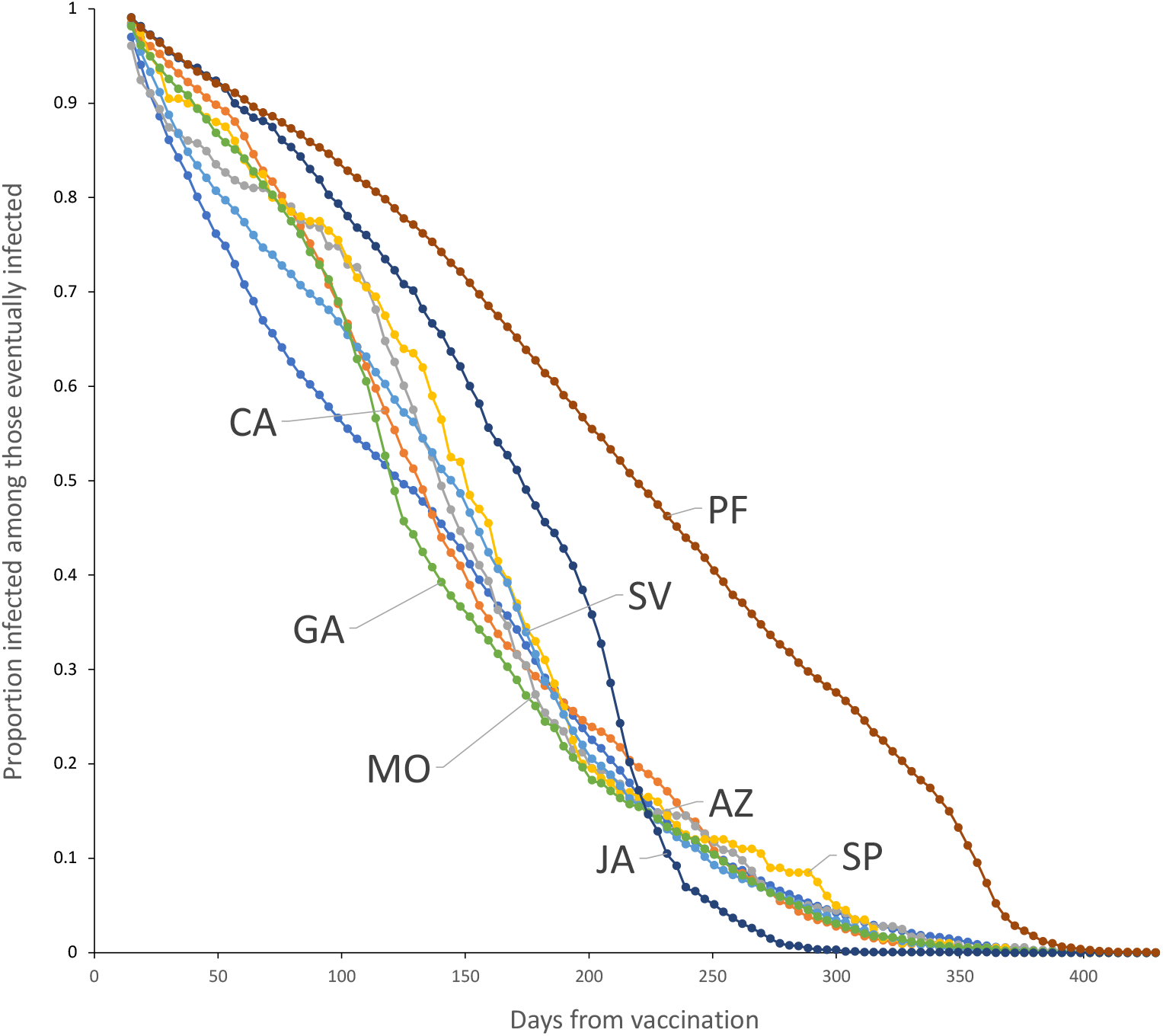
Survival curves for vaccines with more than 100 observations.

**Figure 4:**
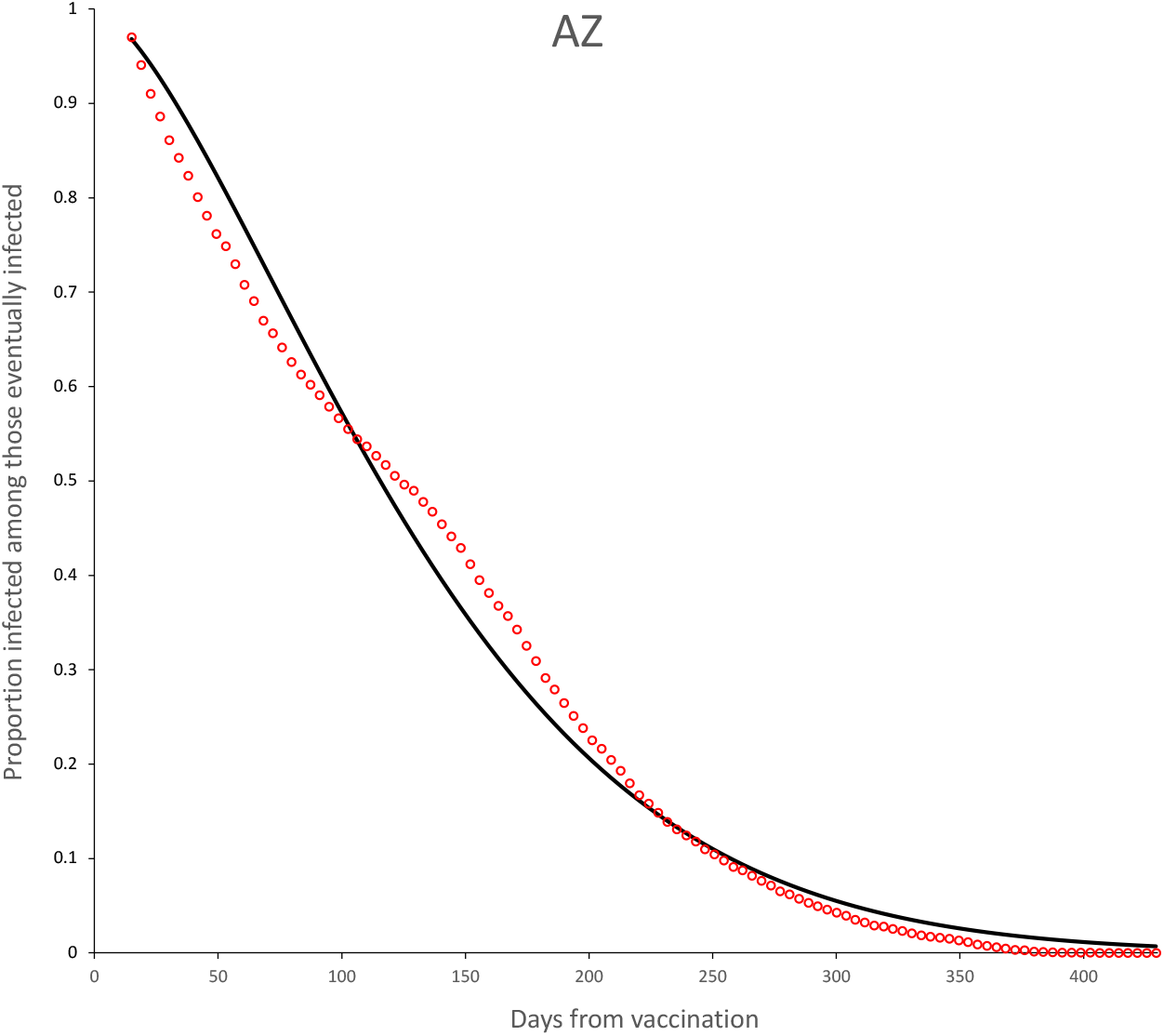
Survival curves for AZ vaccine. Continuous line is the adjusted Weibull survival model (see parameters in Table 5.

**Figure 5:**
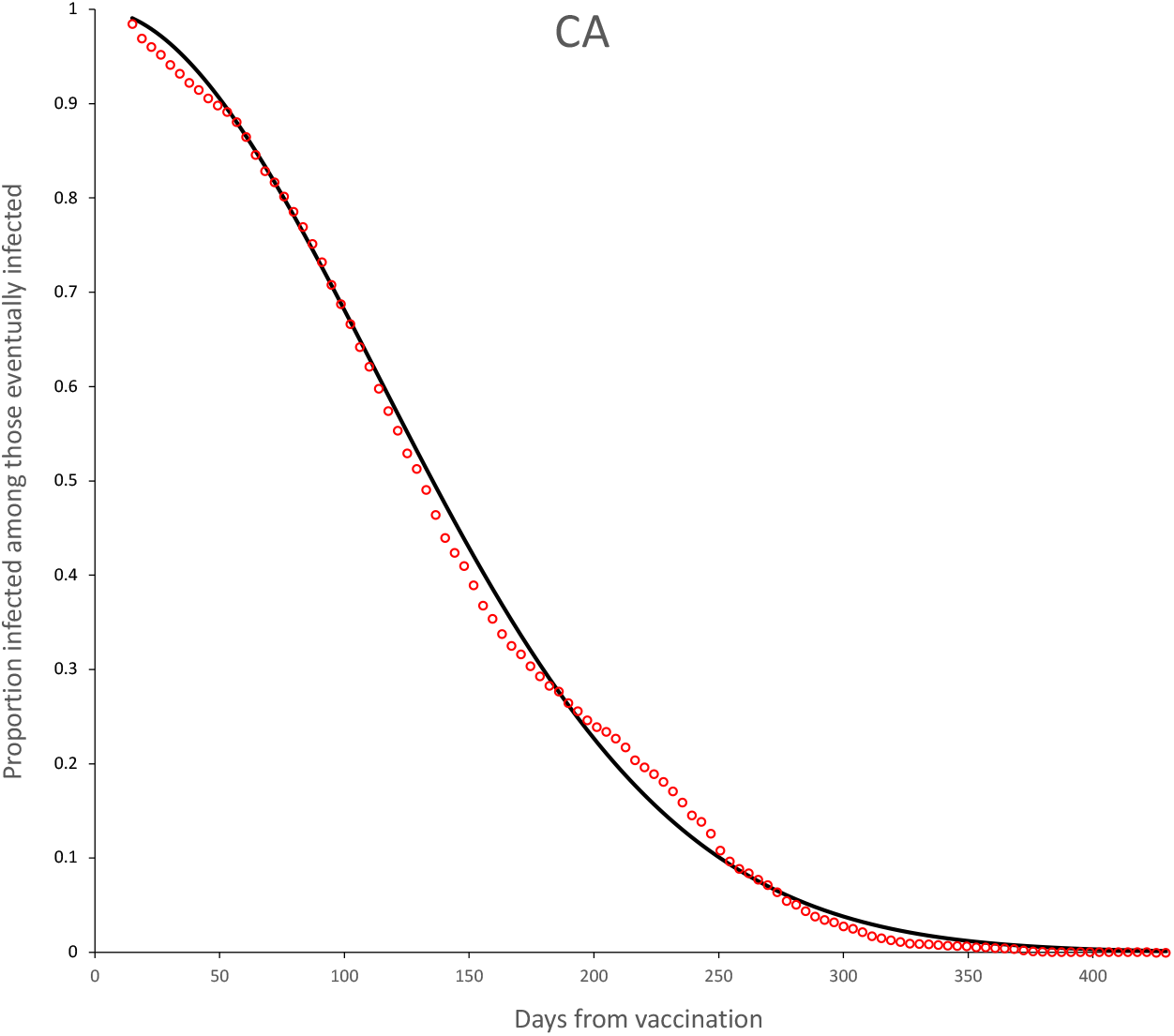
Survival curves for CA vaccine. Continuous line is the adjusted Weibull survival model (see parameters in Table 5.

**Figure 6:**
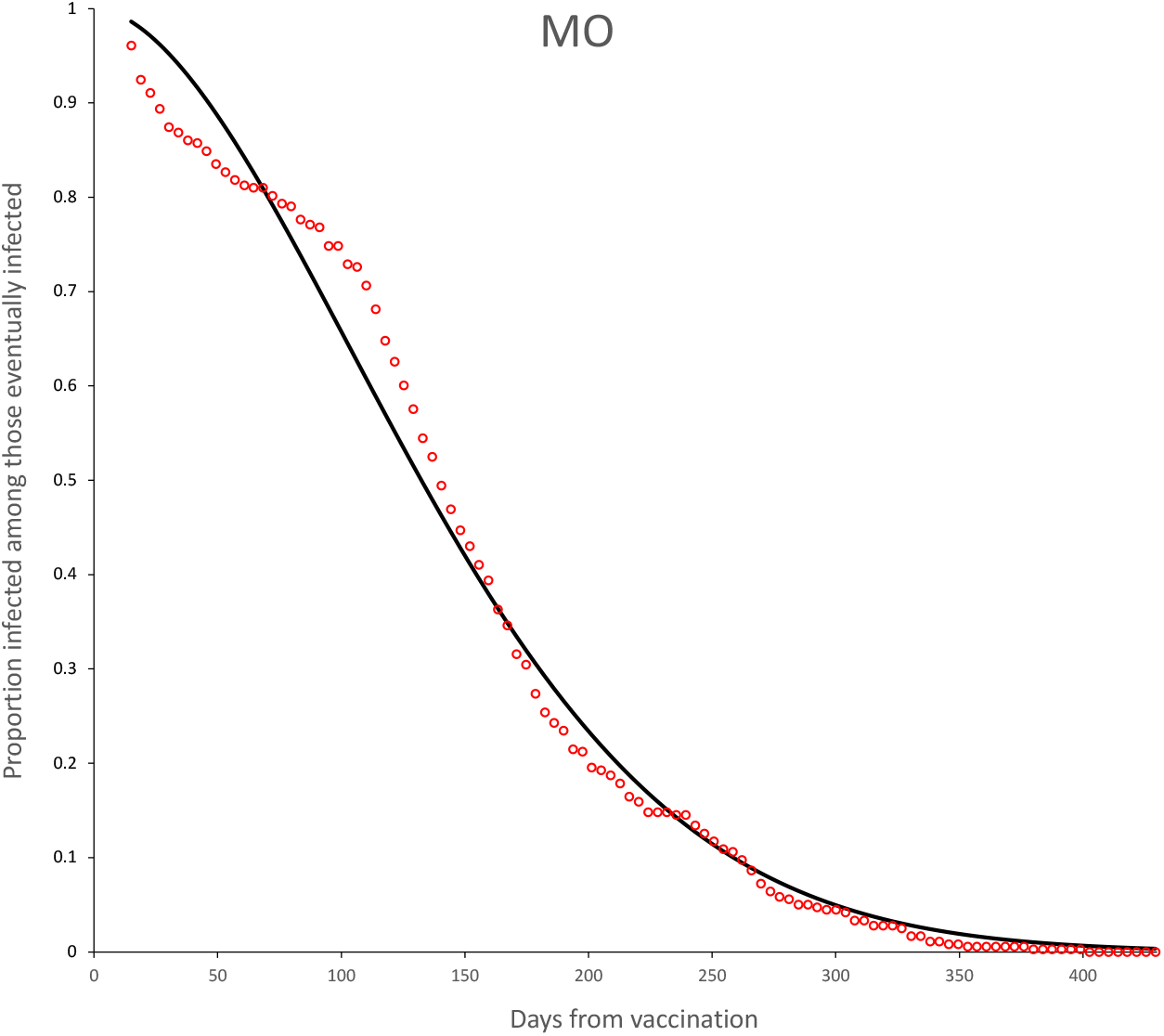
Survival curves for MO vaccine. Continuous line is the adjusted Weibull survival model (see parameters in Table 5.

**Figure 7:**
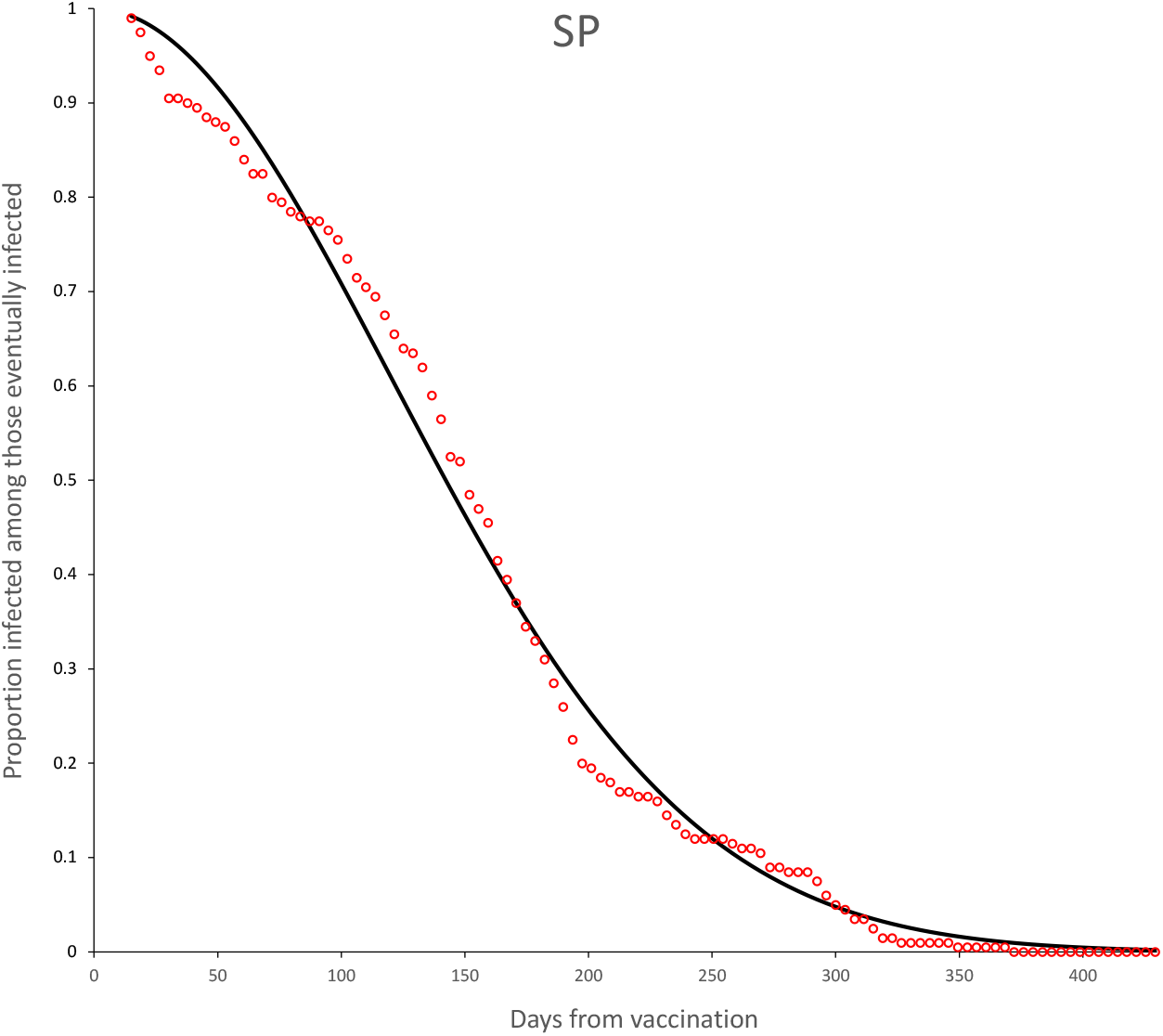
Survival curves for SP vaccine. Continuous line is the adjusted Weibull survival model (see parameters in Table 5.

**Figure 8:**
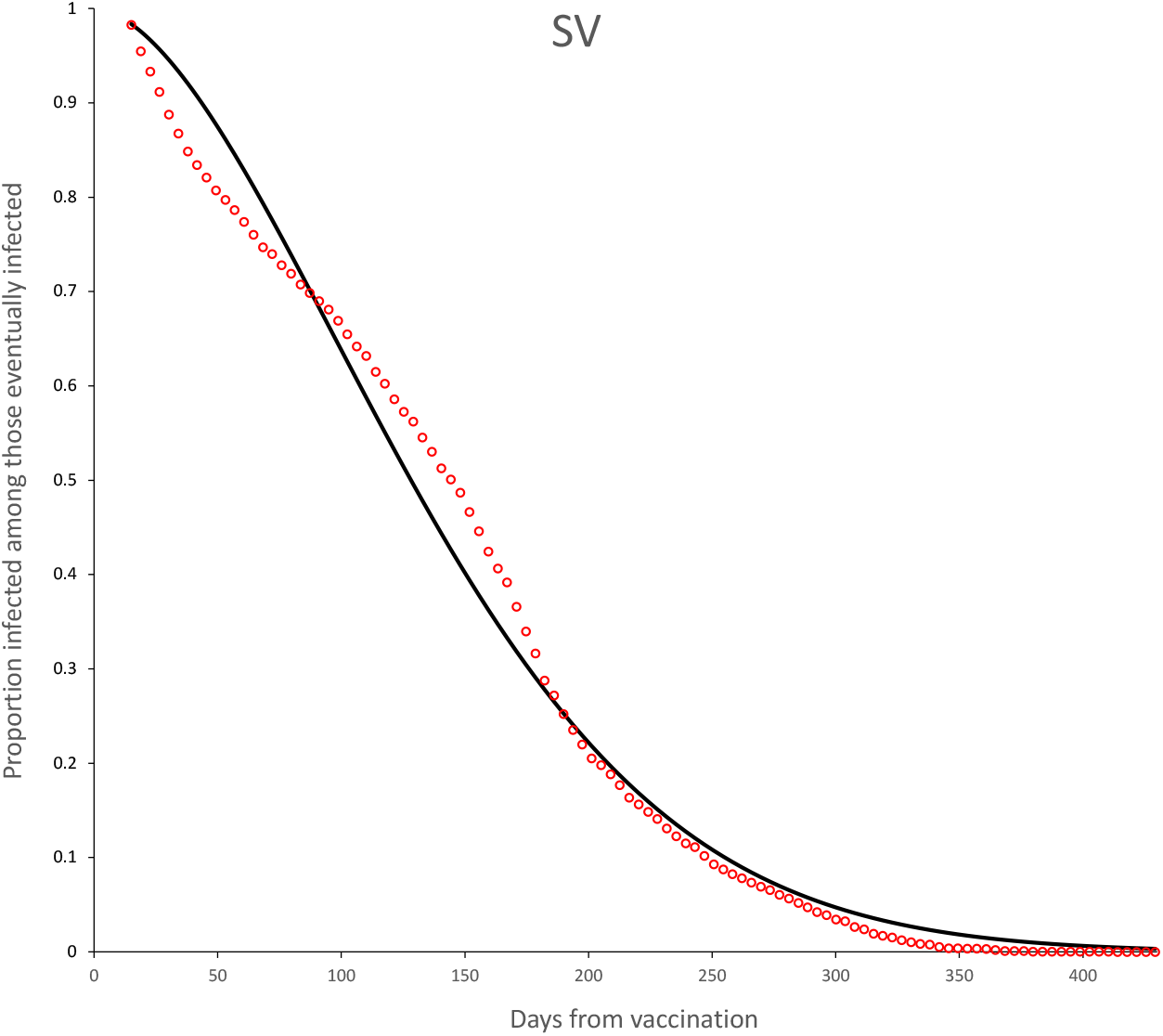
Survival curves for SV vaccine. Continuous line is the adjusted Weibull survival model (see parameters in Table 5.

**Figure 9:**
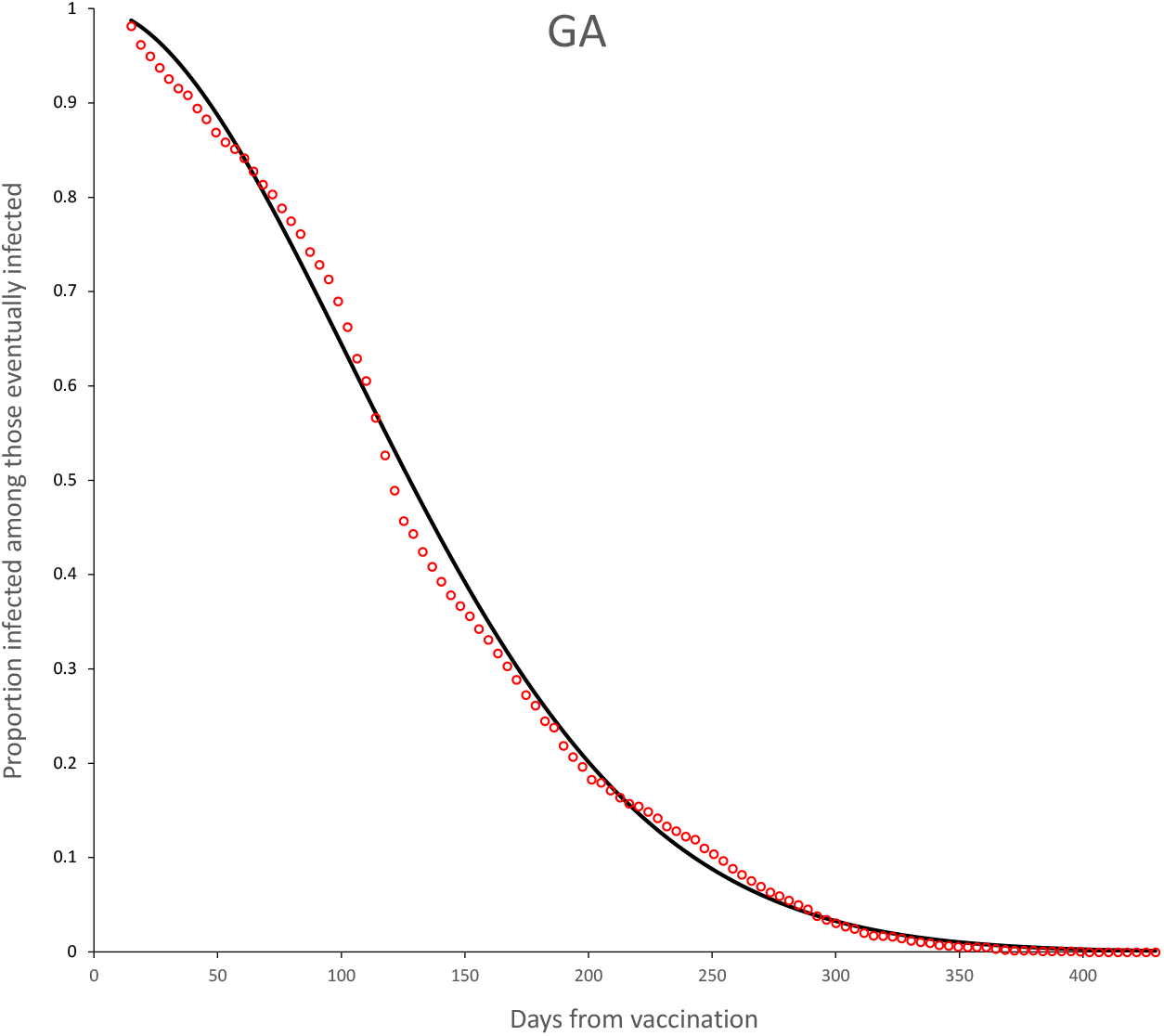
Survival curves for GA vaccine. Continuous line is the adjusted Weibull survival model (see parameters in Table 5.

**Figure 10:**
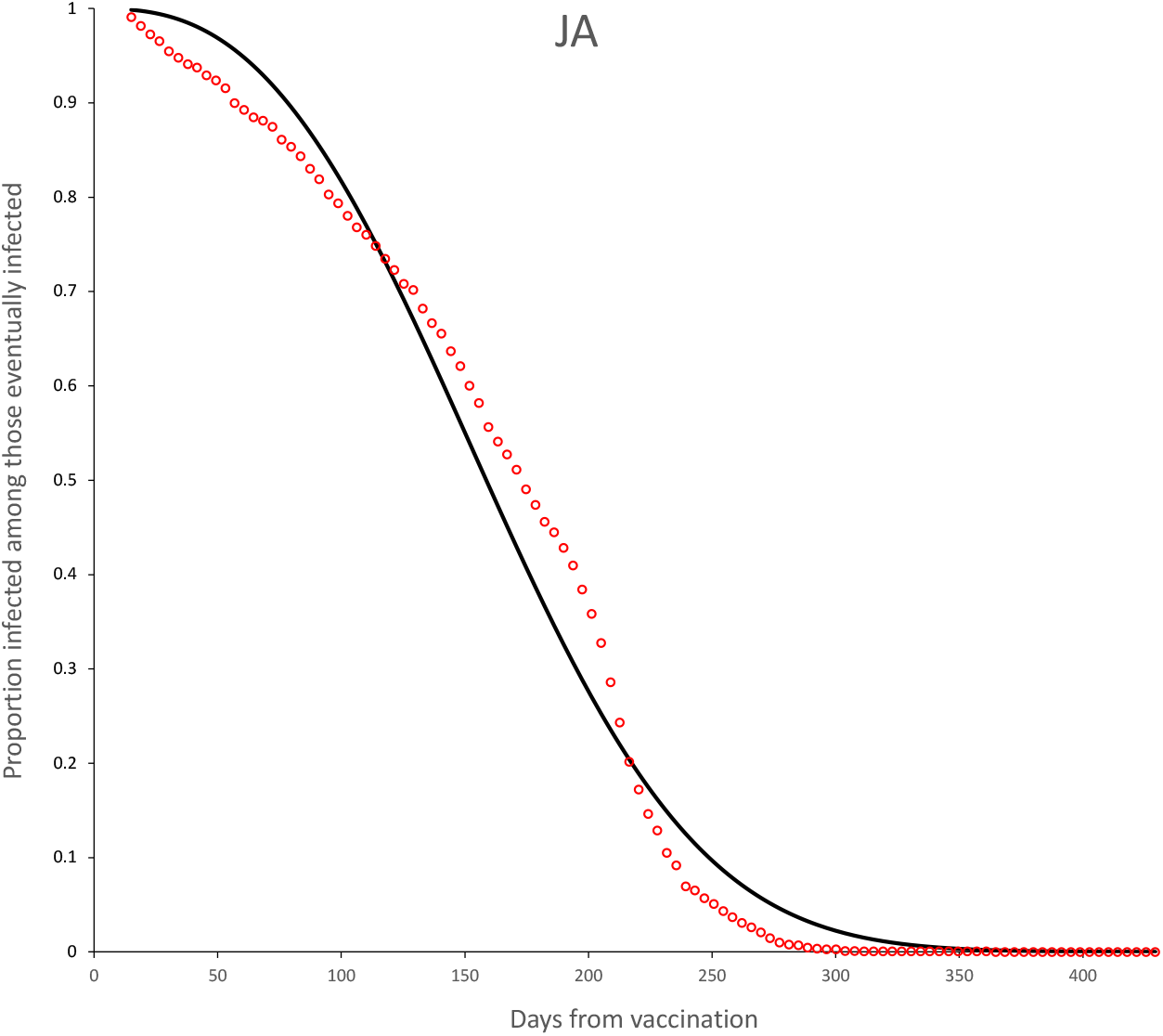
Survival curves for JA vaccine. Continuous line is the adjusted Weibull survival model (see parameters in Table 5.

**Figure 11:**
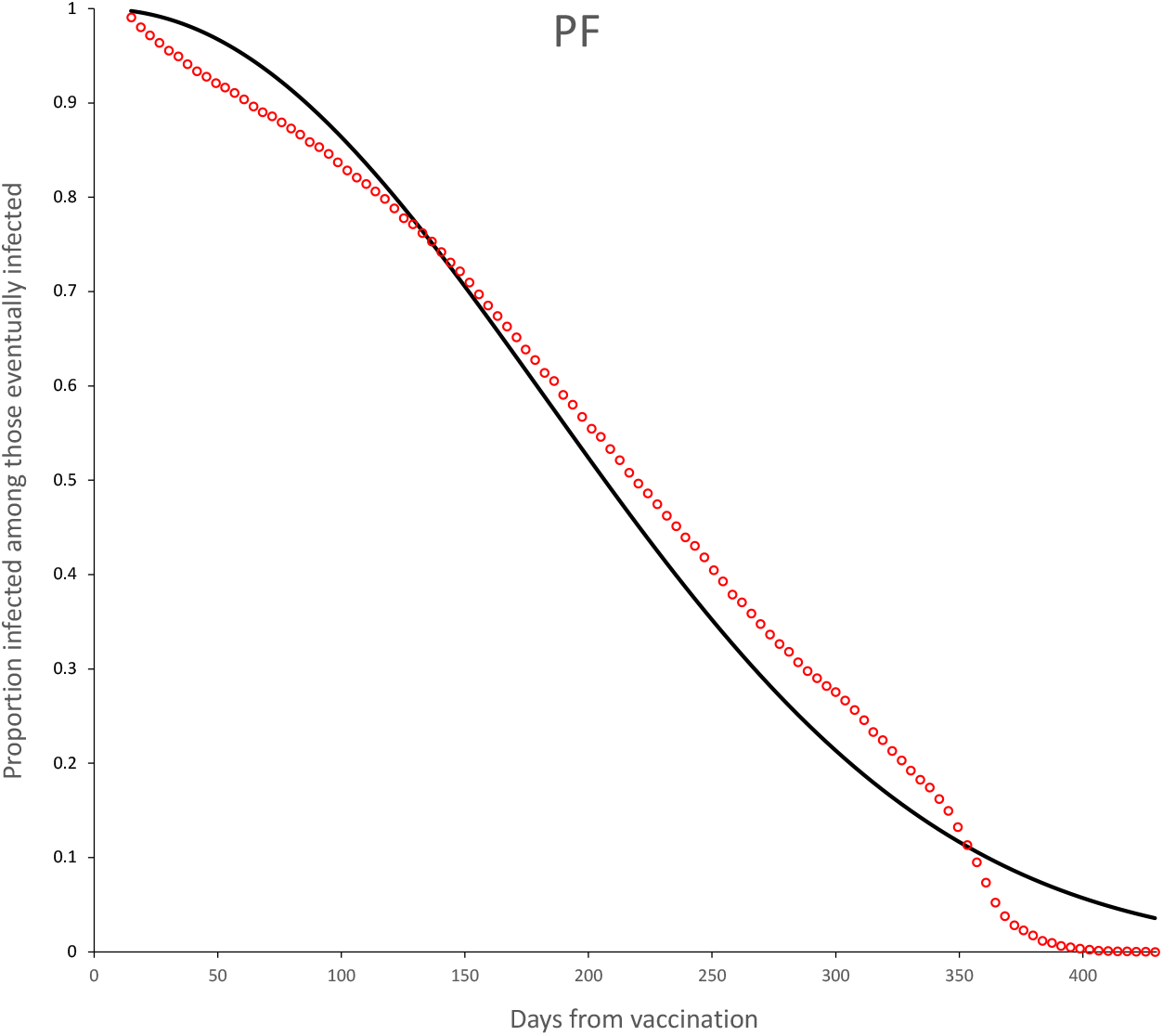
Survival curves for PF vaccine. Continuous line is the adjusted Weibull survival model (see parameters in Table 5.

**Figure 12:**
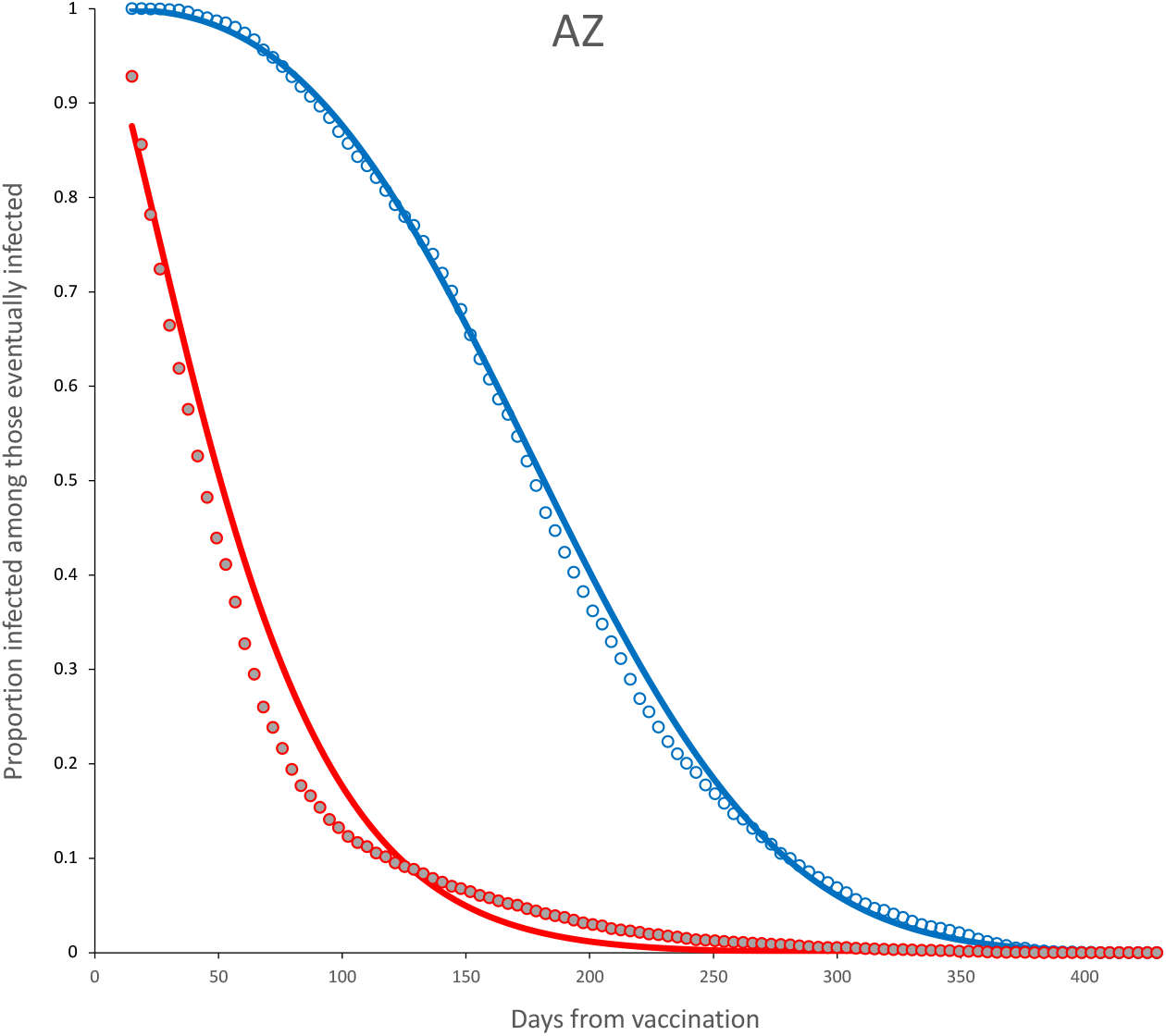
Survival curves and adjusted model for incomplete (red) and complete (blue) dose for AztraZeneca vaccine. Parameter estimates are shown in Table 6.

**Figure 13:**
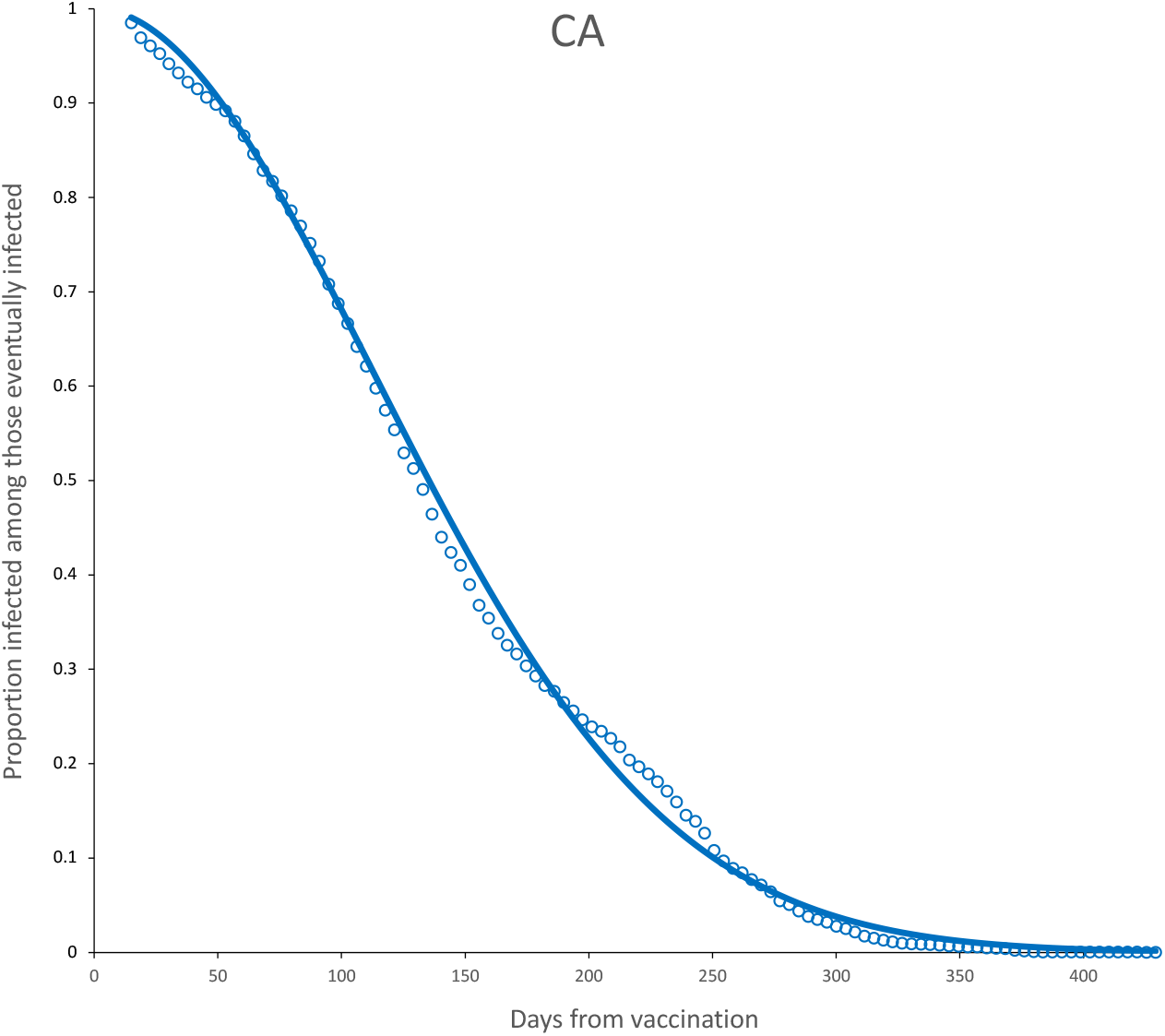
Survival curves and adjusted model for CA vaccine. Parameter estimates are shown in Table 6.

**Figure 14:**
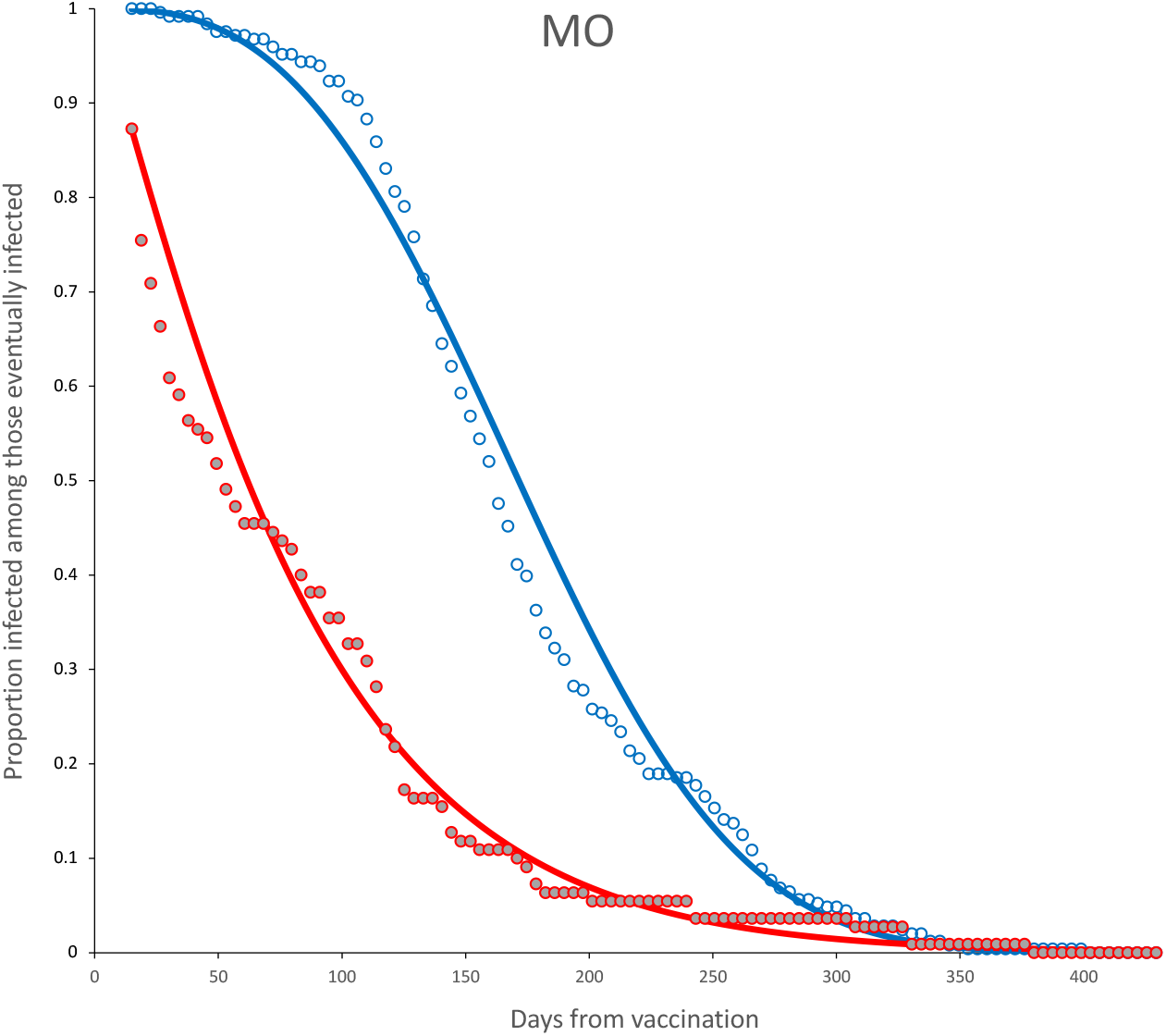
Survival curves and adjusted model for incomplete (red) and complete (blue) dose for MO vaccine. Parameter estimates are shown in Table 6.

**Figure 15:**
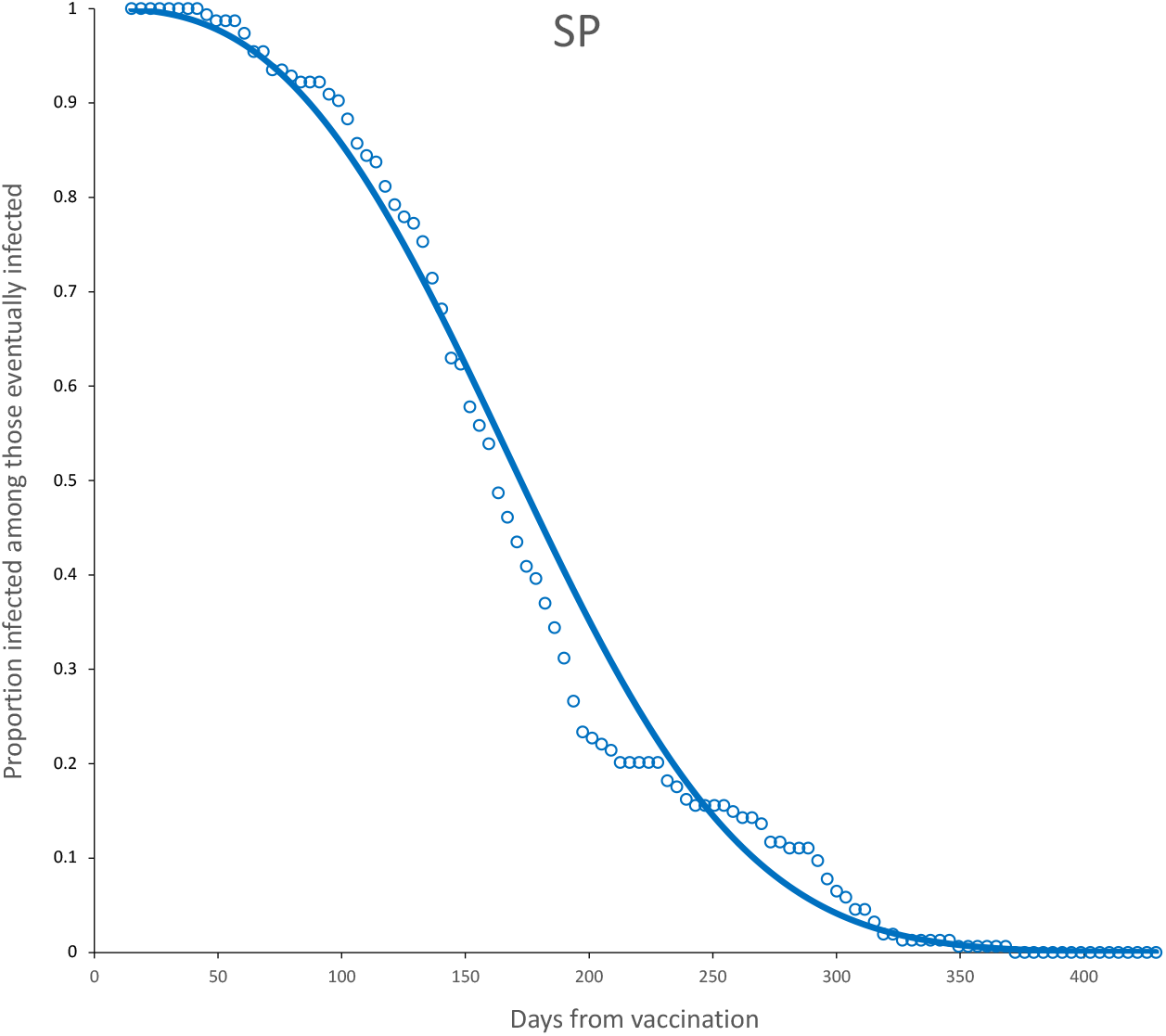
Survival curves and adjusted model for complete (blue) dose for SP vaccine. Parameter estimates are shown in Table 6.

**Figure 16:**
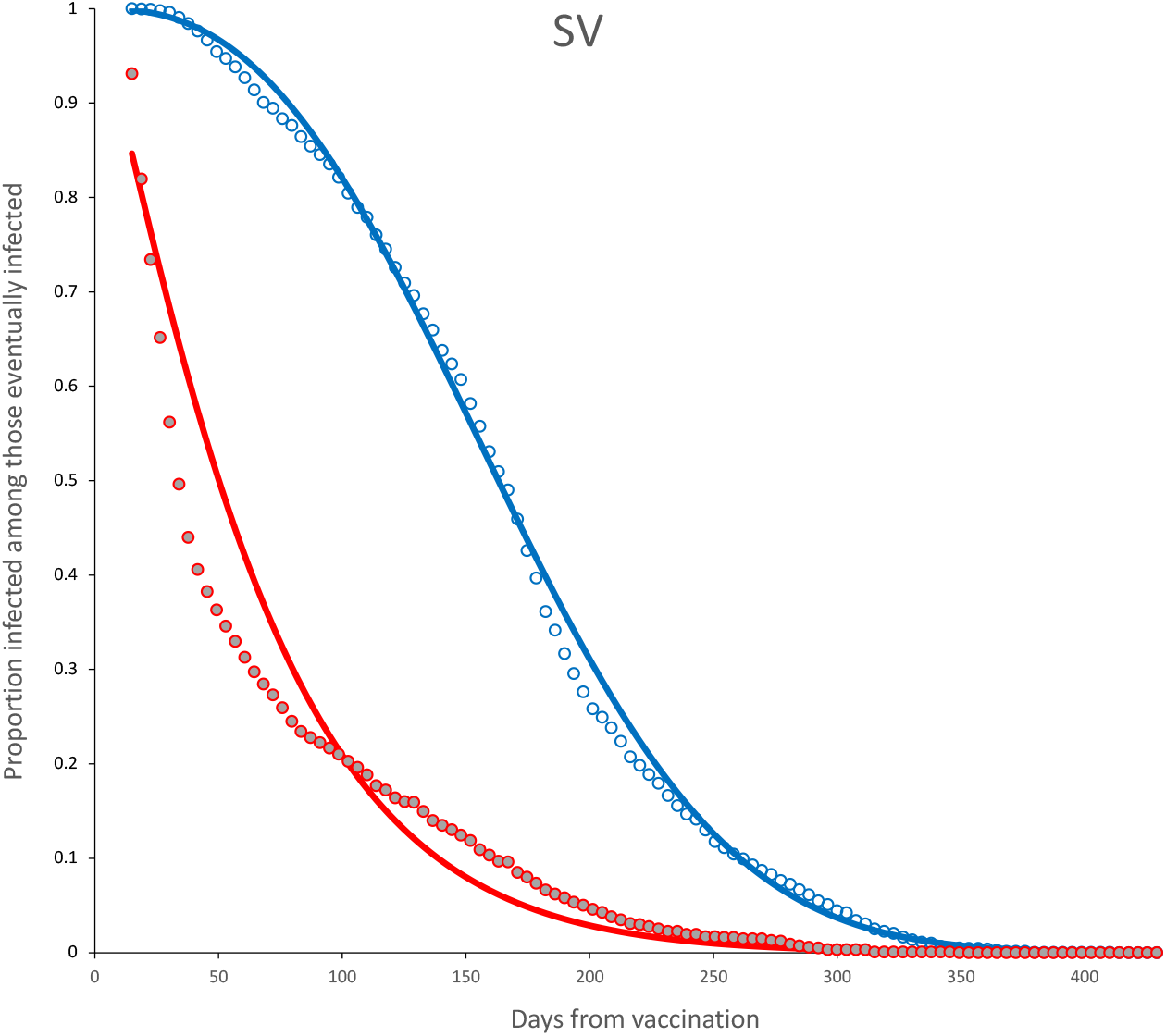
Survival curves and adjusted model for incomplete (red) and complete (blue) dose for SV vaccine. Parameter estimates are shown in Table 6.

**Figure 17:**
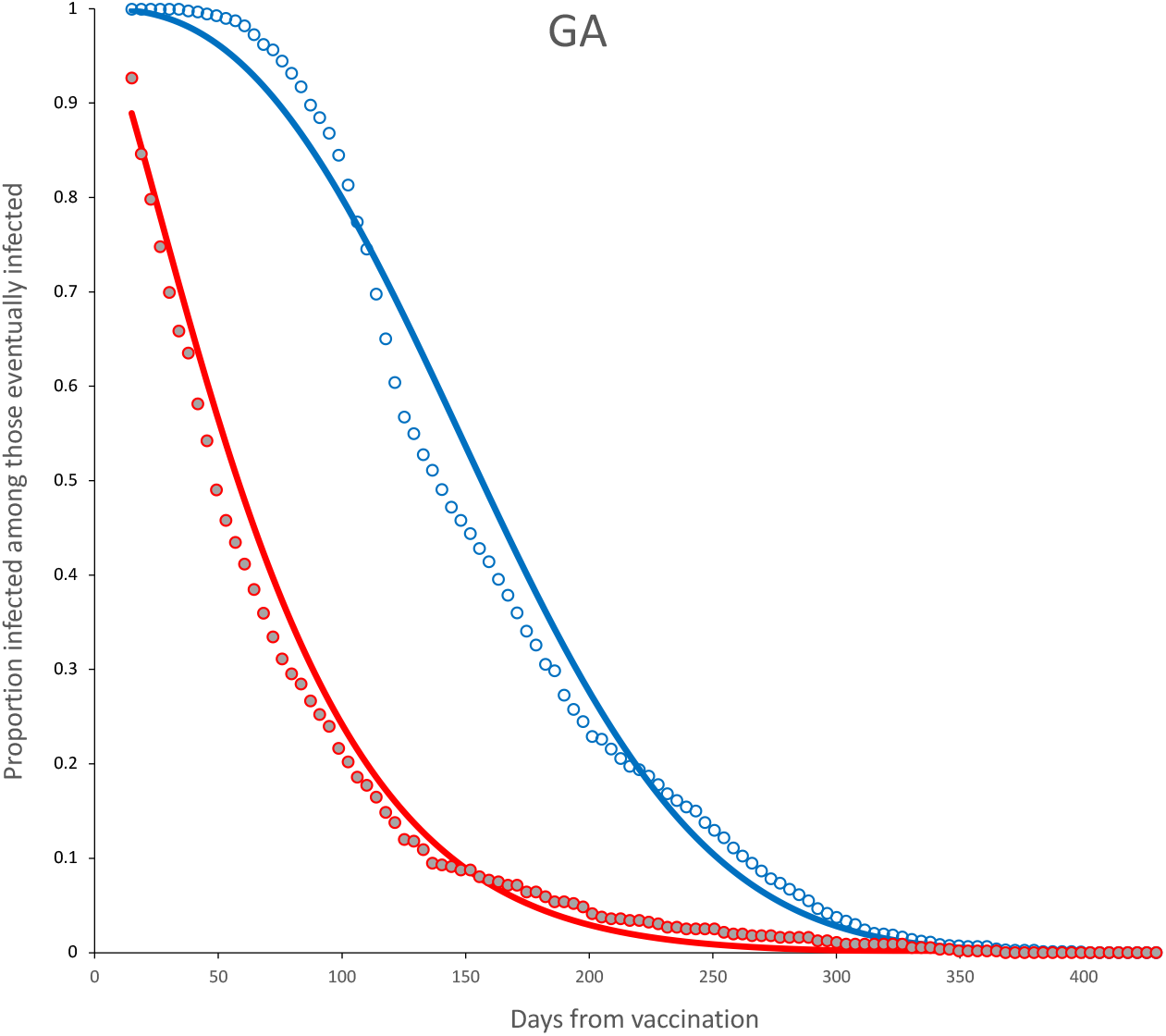
Survival curves and adjusted model for incomplete (red) and complete (blue) dose for GA vaccine. Parameter estimates are shown in Table 6.

**Figure 18:**
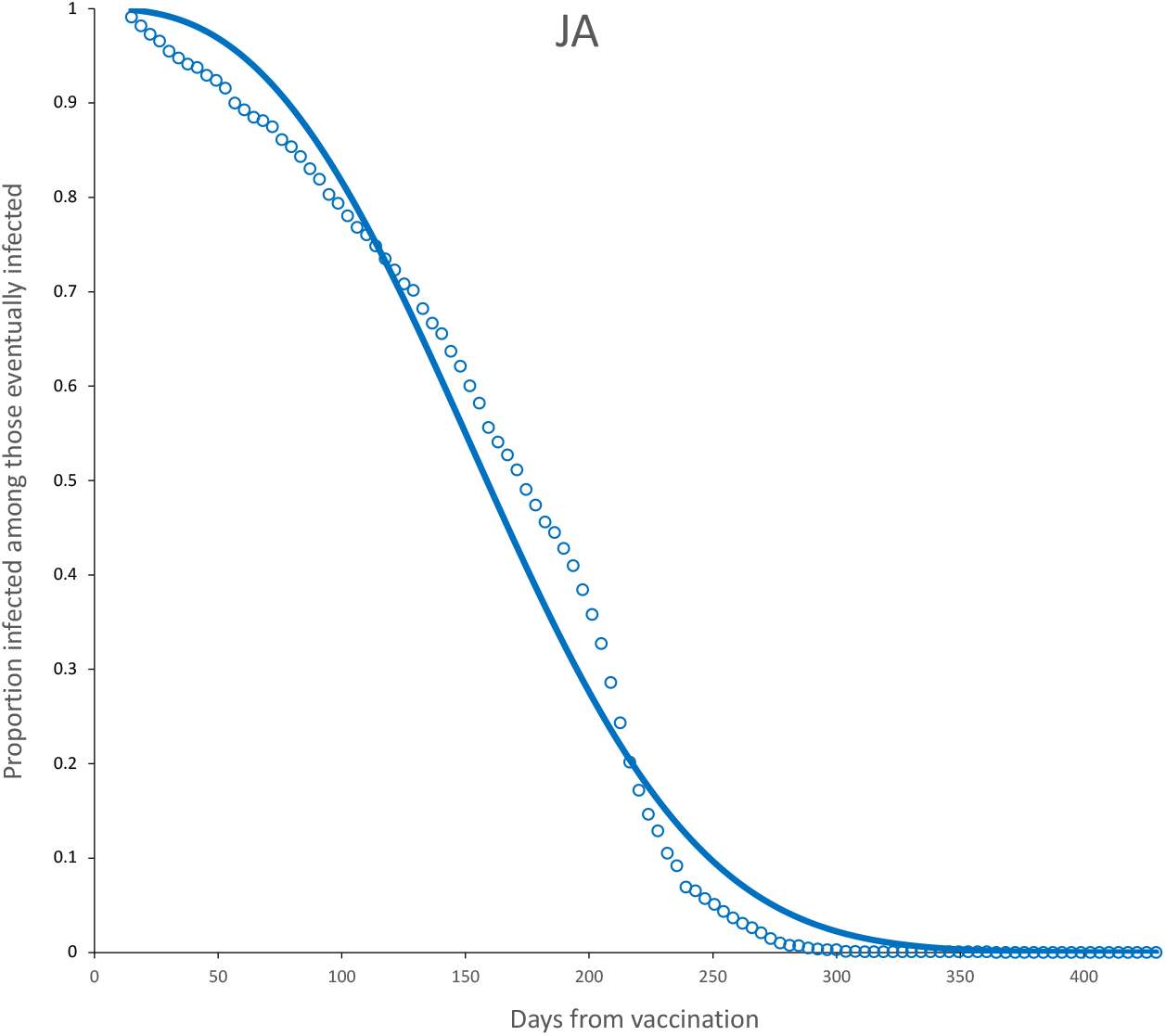
Survival curves and adjusted model for complete (blue) dose for JA vaccine. Parameter estimates are shown in Table 6.

**Figure 19:**
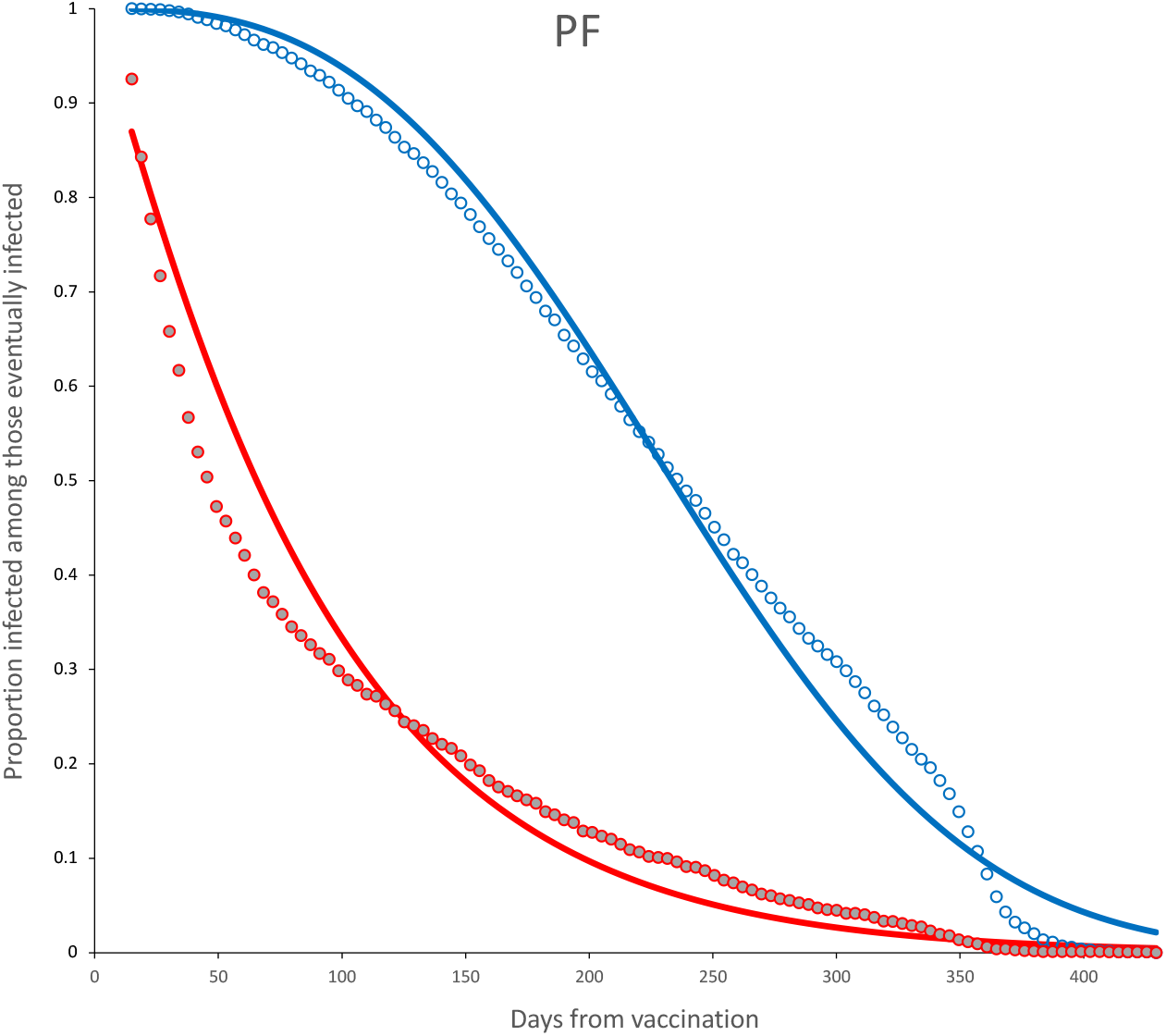
Survival curves and adjusted model for incomplete (red) and complete (blue) dose for PF vaccine. Parameter estimates are shown in Table 6.

**Figure 20:**
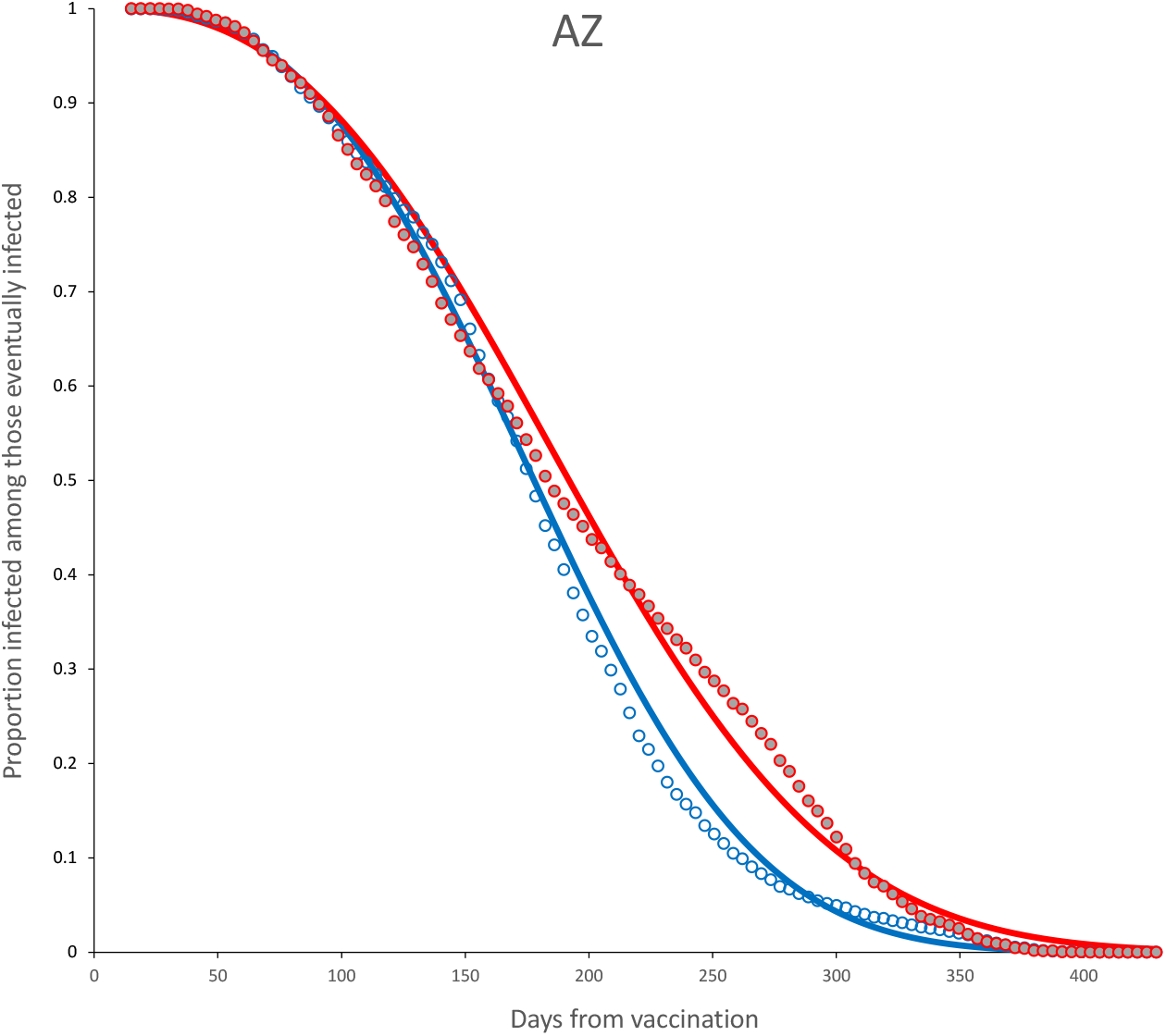
Survival curves and adjusted model for complete dose, age < 60 (blue) and age ≥ 60 (red) for AZ vaccine. Parameter estimates are shown in Table 7.

**Figure 21:**
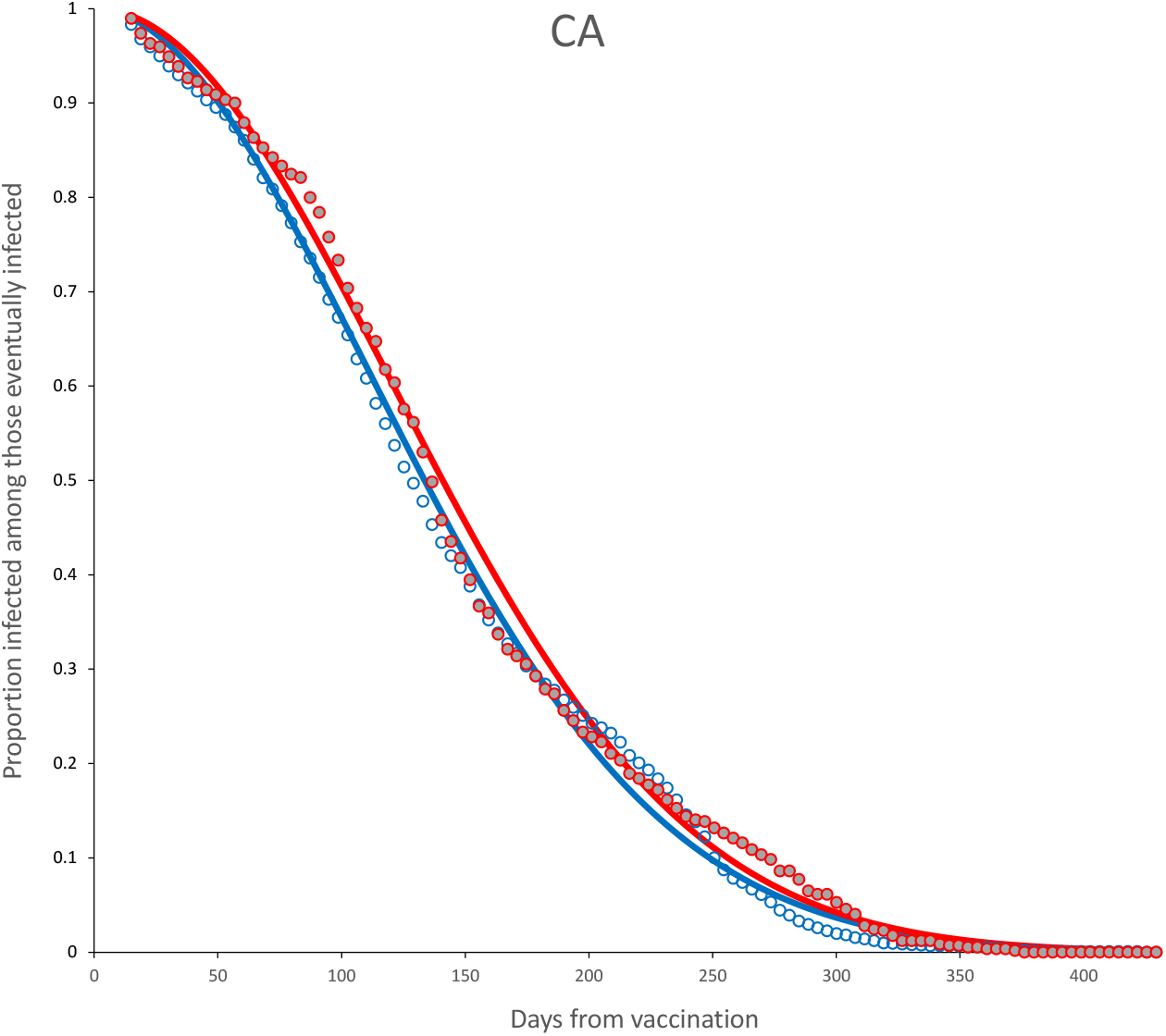
Survival curves and adjusted model for complete dose, age < 60 (blue) and age ≥ 60 (red) for CA vaccine. Parameter estimates are shown in Table 7.

**Figure 22:**
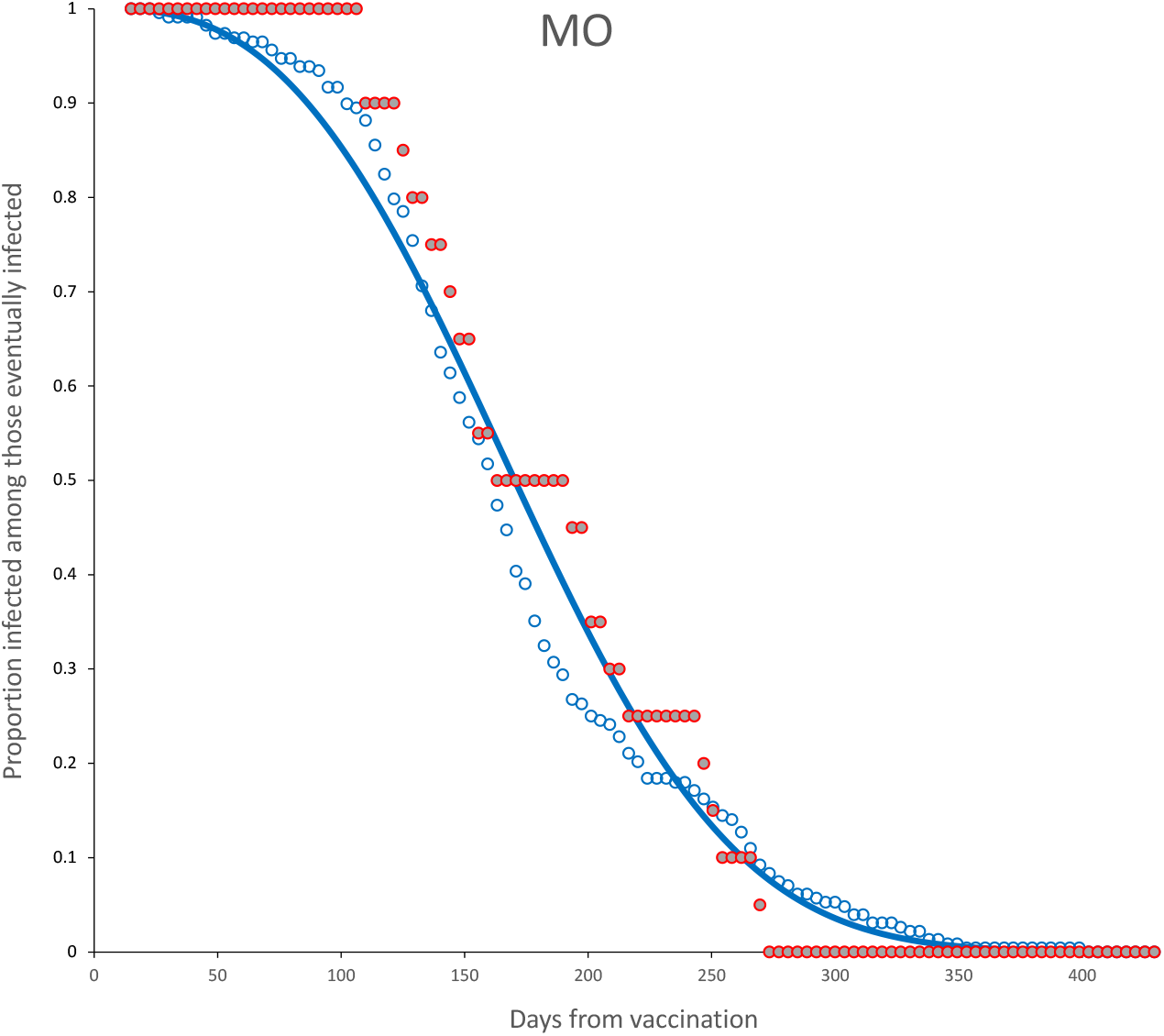
Survival curves and adjusted model for complete dose, age < 60 (blue) for MO vaccine. No model was adjusted for age ≥ 60. Parameter estimates are shown in Table 7.

**Figure 23:**
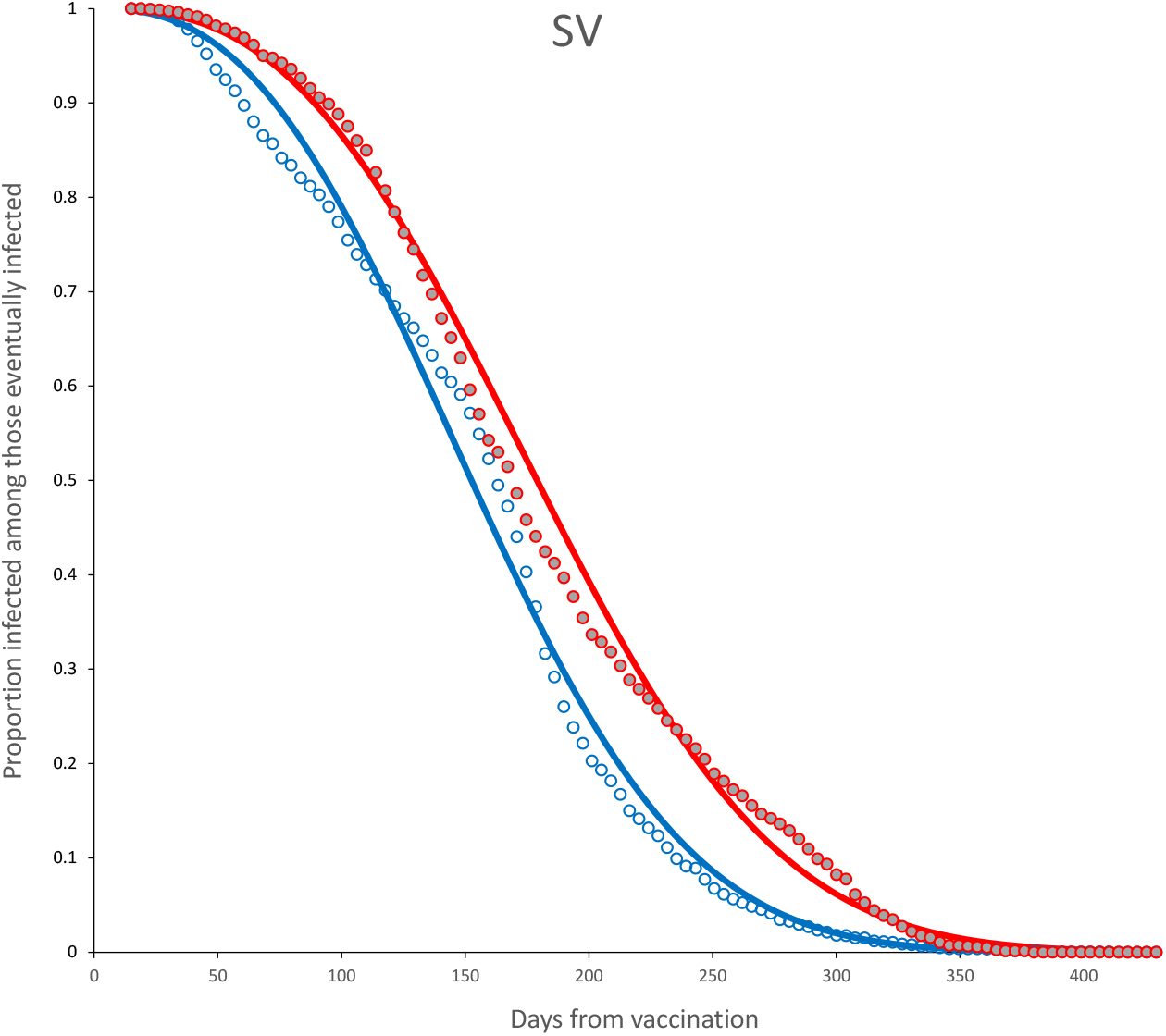
Survival curves and adjusted model for complete dose, age < 60 (blue) and age ≥ 60 (red) for SV vaccine. Parameter estimates are shown in Table 7.

**Figure 24:**
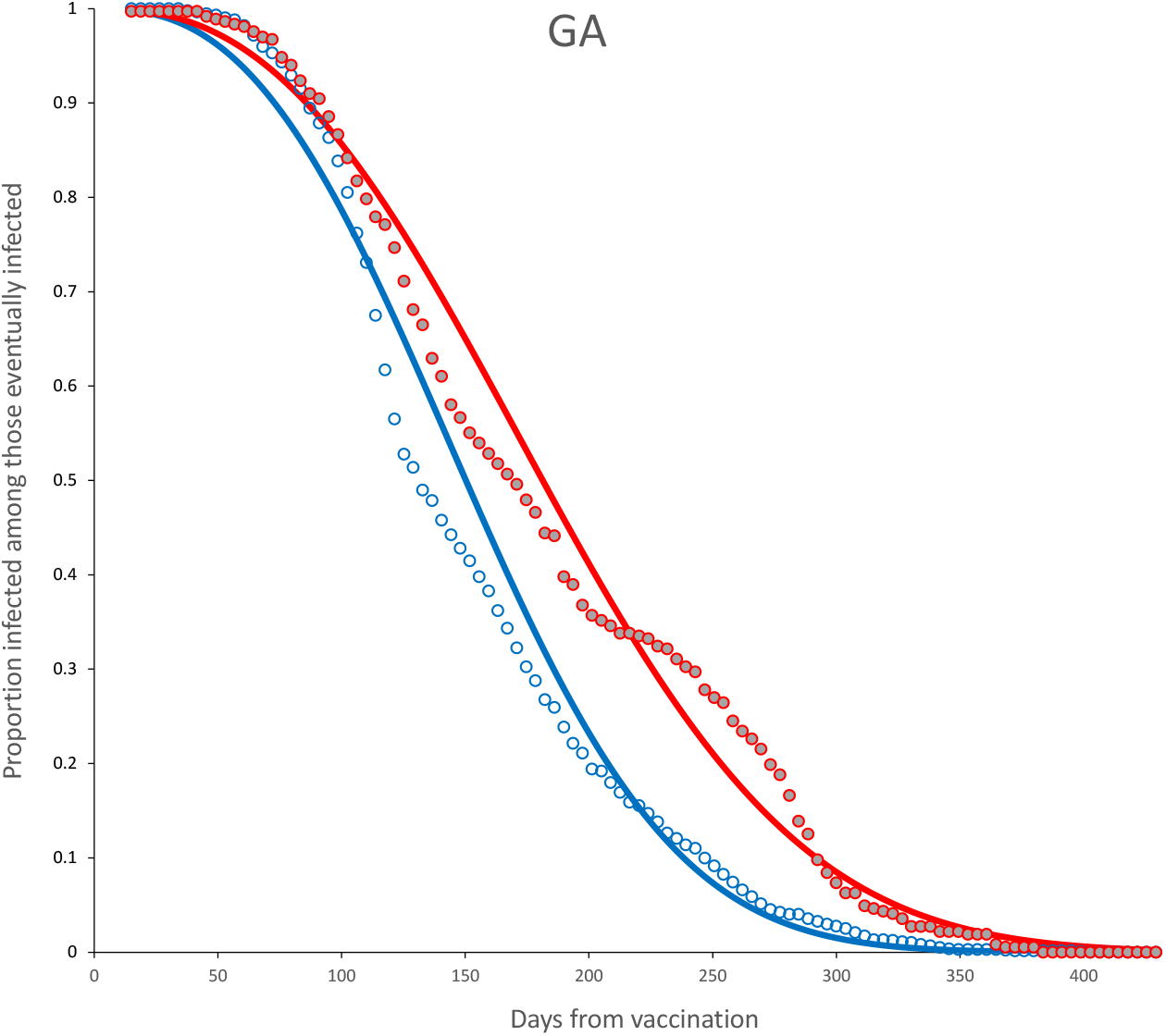
Survival curves and adjusted model for complete dose, age < 60 (blue) and age ≥ 60 (red) for GA vaccine. Parameter estimates are shown in Table 7.

**Figure 25:**
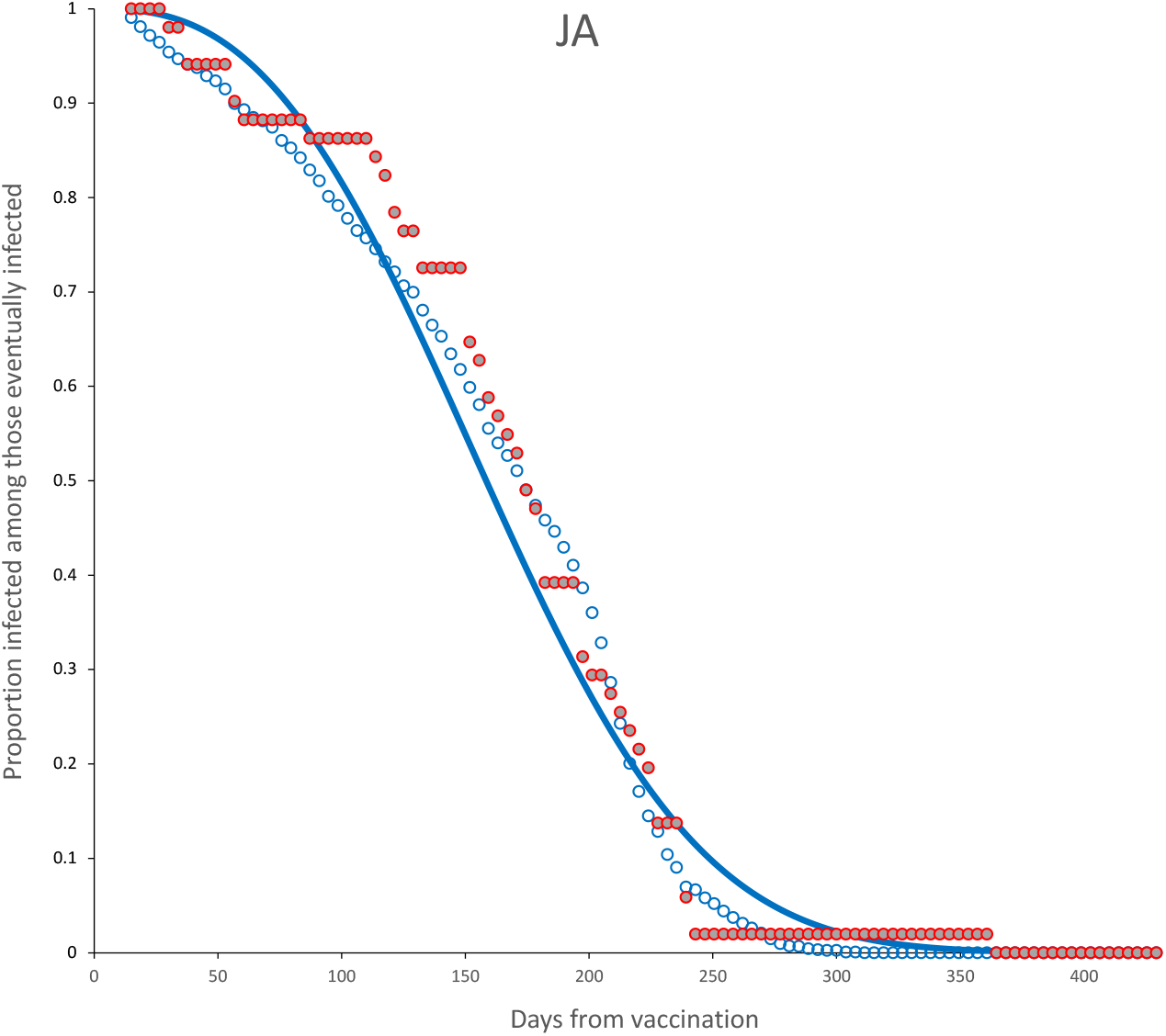
Survival curves and adjusted model for complete dose, age < 60 (blue) for JA vaccine. No model was adjusted for age ≥ 60. Parameter estimates are shown in Table 7.

**Figure 26:**
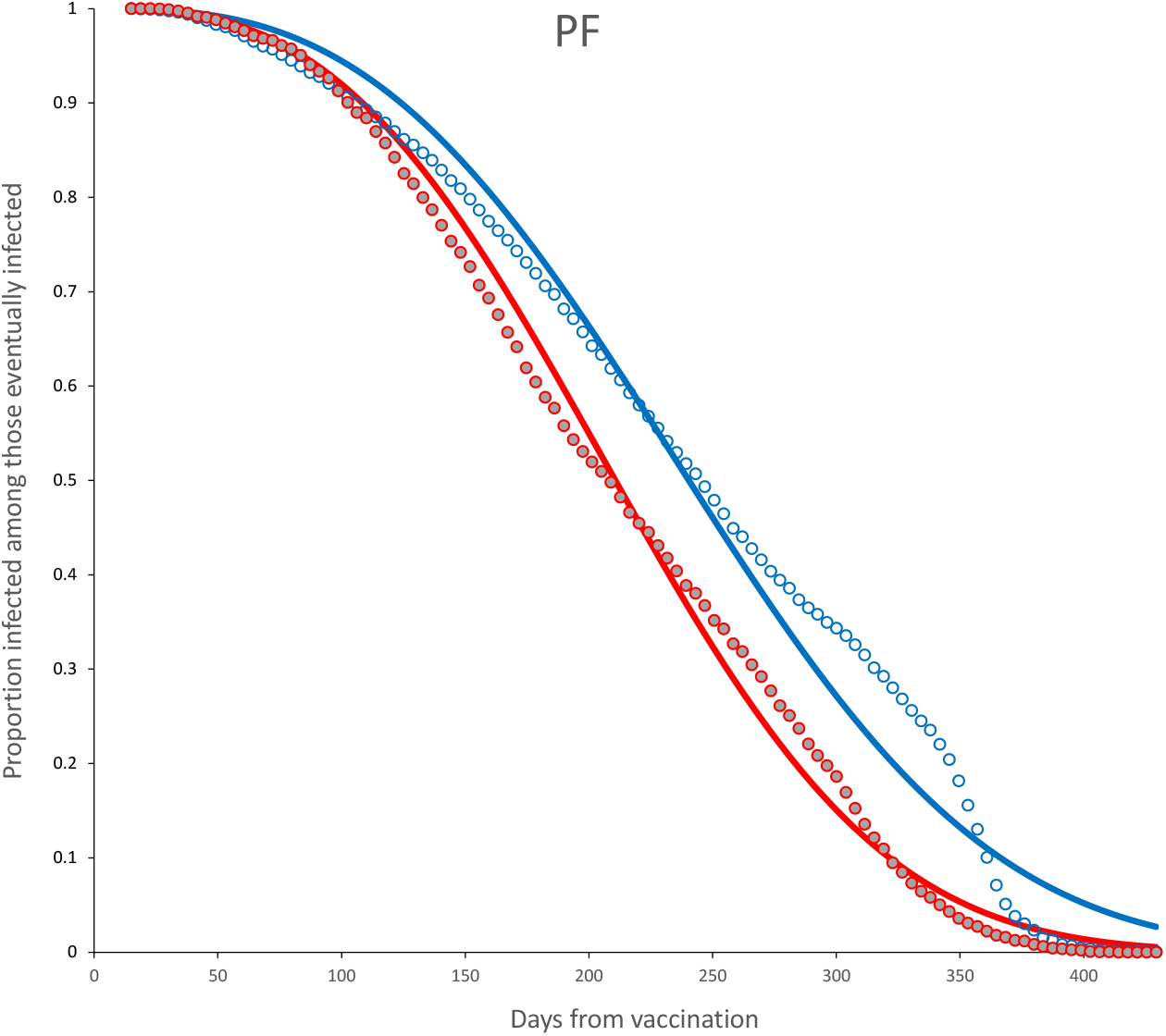
Survival curves and adjusted model for complete dose, age < 60 (blue) and age ≥ 60 (red) for PF vaccine. Parameter estimates are shown in Table 7.

**Figure 27:**
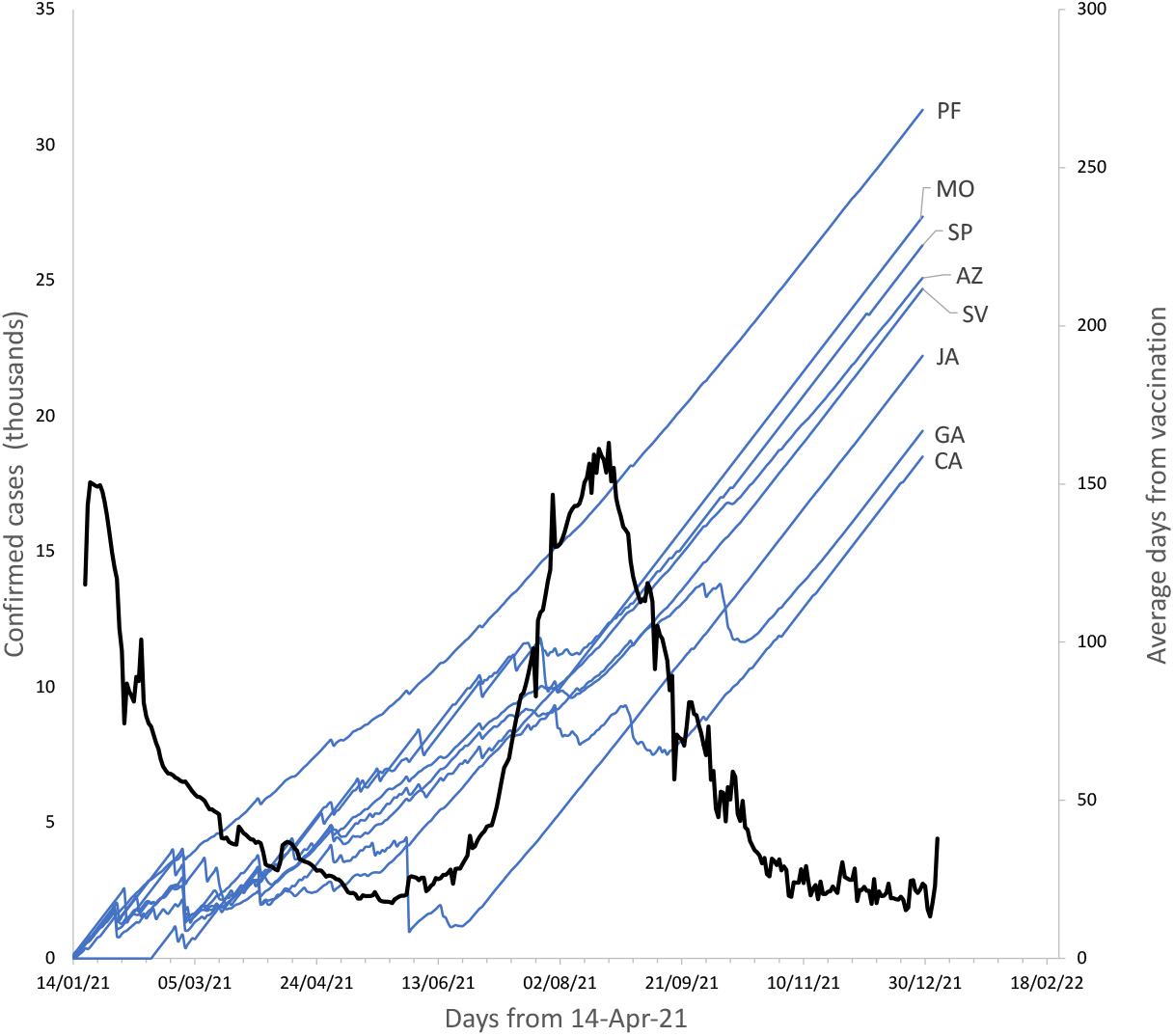
Deployment of the different vaccines through from one year after beginning of mass vaccination. The thick, black line indicates the evolution of the confirmed SARS-CoV-2 cases in Mexico (Dong et al., 2020), as a surrogate of infection pressure (left axis). The rest of the lines show, for each vaccine, the evolution of the mean *vaccine age* (right axis).

## Bibliography

Afshar, Z. M., Barary, M., Hosseinzadeh, R., Alijanpour, A., Hosseinzadeh, D., Ebrahimpour, S., Nazary, K., Sio, T. T., Sullman, M. J. M., Carson-Chahhoud, K., & Babazadeh, A. (). Breakthrough sars-cov-2 infections after vaccination: a critical review. HUMAN VACCINES & IMMUNOTHERAPEUTICS,. doi:10.1080/21645515.2022.2051412.

Andrews, N., Tessier, E., Stowe, J., Gower, C., Kirsebom, F., Simmons, R., Gallagher, E., Thelwall, S., Groves, N., Dabrera, G., Myers, R., Campbell, C. N. J., Amirthalingam, G., Edmunds, M., Zambon, M., Brown, K., Hopkins, S., Chand, M., Ladhani, S. N., Ramsay, M., & Bernal, J. L. (2022). Duration of protection against mild and severe disease by covid-19 vaccines. NEW ENGLAND JOURNAL OF MEDICINE, 386, 340–350. doi:10.1056/NEJMoa2115481.

Bedston, S., Akbari, A., Jarvis, C. I., Lowthian, E., Torabi, F., North, L., Lyons, J., Perry, M., Griffiths, L. J., Owen, R. K., Beggs, J., Chuter, A., Bradley, D. T., de Lusignan, S., Fry, R., Hobbs, F. D. R., Hollinghurst, J., Katikireddi, S. V., Murphy, S., O’Reily, D., Robertson, C., Shi, T., Tsang, R. S. M., Sheikh, A., & Lyons, R. A. (2022). Covid-19 vaccine uptake, effectiveness, and waning in 82,959 health care workers: A national prospective cohort study in wales. VACCINE, 40, 1180–1189. doi:10.1016/j.vaccine.2021.11.061.

Berar-Yanay, N., Freiman, S., Shapira, M., Saffoury, A., Elemy, A., Hamze, M., Elhaj, M., Zaher, M., Matanis, L., & Armaly, Z. A. (2021). Waning humoral response 3 to 6 months after vaccination with the sars-cov-2 bnt162b2 mrna vaccine in dialysis patients. JOURNAL OF CLINICAL MEDICINE, 11, 64.

Blower, S., & McLean, A. (1994). Prophylactic vaccines, risk behavior change, and the probability of eradicating hiv in san francisco. SCIENCE, 265, 1451–1454. doi:10.1126/science.8073289.

Coppeta, L., Ferrari, C., Somma, G., Mazza, A., D’Ancona, U., Marcuccilli, F., Grelli, S., Aurilio, M. T., Pietroiusti, A., Magrini, A., & Rizza, S. (2022). Reduced titers of circulating anti-sars-cov-2 antibodies and risk of covid-19 infection in healthcare workers during the nine months after immunization with the bnt162b2 mrna vaccine. VACCINES, 10. doi:10.3390/vaccines10020141.

Dong, E., Du, H., & Gardner, L. (2020). An interactive web-based dashboard to track covid-19 in real time. LANCET INFECTIOUS DISEASES, 20, 533–534. doi:10.1016/S1473-3099(20)30120-1.

Ferdinands, J. M., Rao, S., Dixon, B. E., Mitchell, P. K., DeSilva, M. B., Irving, S. A., Lewis, N., Natarajan, K., Stenehjem, E., Grannis, S. J., Han, J., McEvoy, C., Ong, T. C., Naleway, A. L., Reese, S. E., Embi, P. J., Dascomb, K., Klein, N. P., Griggs, E. P., Konatham, D., Kharbanda, A. B., Yang, D.-H., Fadel, W. F., Grisel, N., Goddard, K., Patel, P., Liao, I.-C., Birch, R., Valvi, N. R., Reynolds, S., Arndorfer, J., Zerbo, O., Dickerson, M., Murthy, K., Williams, J., Bozio, C. H., Blanton, L., Verani, J. R., Schrag, S. J., Dalton, A. F., Wondimu, M. H., Link-Gelles, R., Azziz-Baumgartner, E., Barron, M. A., Gaglani, M., Thompson, M. G., & Fireman, B. (2022). Waning 2-dose and 3-dose effectiveness of mrna vaccines against covid-19-associated emergency department and urgent care encounters and hospitalizations among adults during periods of delta and omicron variant predominance - vision network, 10 states, august 2021-january 2022. MMWR-MORBIDITY AND MORTALITY WEEKLY REPORT, 71, 255–263.

Fukushima, W., & Hirota, Y. (2017). Basic principles of test-negative design in evaluating influenza vaccine effectiveness. VACCINE, 35, 4796–4800. URL: https://www.sciencedirect.com/science/article/pii/S0264410X1730899X. doi: https://doi.org/10.1016/j.vaccine.2017.07.003.

Goldberg, Y., Mandel, M., Bar-On, Y. M., Bodenheimer, O., Freedman, L., Haas, E. J., Milo, R., Alroy-Preis, S., Ash, N., & Huppert, A. (2021). Waning immunity after the bnt162b2 vaccine in israel. NEW ENGLAND JOURNAL OF MEDICINE, 385, E85. doi:10.1056/NEJMoa2114228.

Housset, P., Kubab, S., Pardon, A., Vittoz, N., Bozman, D.-F., Hanafi, L., Caudwell, V., & Faucon, A.-L. (2022). Waning but persistent humoral response 6 months after the third dose of the mrna bnt162b2 vaccine in hemodialysis and peritoneal dialysis patients. JOURNAL OF NEPHROLOGY, (pp. 1–3). doi:10.1007/s40620-022-01276-2.

IMSS (2022). Datos abiertos imss. URL: http://datos.imss.gob.mx/.

Johnson, N. L., Kemp, A. W., & Kotz, S. (2005). Univariate discrete distributions volume 444. John Wiley & Sons.

Khoury, J., Najjar-Debbiny, R., Hanna, A., Jabbour, A., Abu Ahmad, Y., Saffuri, A., Abu-Sinni, M., Shkeiri, R., Elemy, A., & Hakim, F. (2021). Covid-19 vaccine-long term immune decline and breakthrough infections. VACCINE, 39, 6984–6989. doi:10.1016/j.vaccine.2021.10.038.

Kolaric, B., Ambriovic-Ristov, A., Tabain, I., & Vilibic-Cavlek, T. (2021). Waning immunity six months after biontech/pfizer covid-19 vaccination among nursing home residents in zagreb, croatia. CROATIAN MEDICAL JOURNAL, 62, 630–633. doi:10.3325/cmj.2021.62.630.

Kurita, J., Sugawara, T., & Ohkusa, Y. (2022). Waning covid-19 vaccine effectiveness in japan. DRUG DISCOVERIES AND THERAPEUTICS, 16, 30–36. doi:10.5582/ddt.2022.01000.

Leung, K.-M., Elashoff, R. M., & Afifi, A. A. (1997). Censoring issues in survival analysis. ANNUAL REVIEW OF PUBLIC HEALTH, 18, 83–104.

Levin, E. G., Lustig, Y., Cohen, C., Fluss, R., Indenbaum, V., Amit, S., Doolman, R., Asraf, K., Mendelson, E., Ziv, A., Rubin, C., Freedman, L., Kreiss, Y., & Regev-Yochay, G. (2021). Waning immune humoral response to bnt162b2 covid-19 vaccine over 6 months. NEW ENGLAND JOURNAL OF MEDICINE, 385, E84. doi:10.1056/NEJMoa2114583.

Levine-Tiefenbrun, M., Yelin, I., Alapi, H., Herzel, E., Kuint, J., Chodick, G., Gazit, S., Patalon, T., & Kishony, R. (2022). Waning of sars-cov-2 booster viral-load reduction effectiveness. NATURE COMMUNICATIONS, 13. doi:10.1038/s41467-022-28936-y.

Lin, D.-Y., Gu, Y., Zeng, D., Janes, H. E., & Gilbert, P. B. (2022). Evaluating vaccine efficacy against severe acute respiratory syndrome coronavirus 2 infection. CLINICAL INFECTIOUS DISEASES, 74, 544–552. doi:10.1093/cid/ciab630.

Nordstrom, P., Ballin, M., & Nordstrom, A. (2022). Risk of infection, hospitalisation, and death up to 9 months after a second dose of covid-19 vaccine: a retrospective, total population cohort study in sweden. LANCET, 399, 814–823. doi:10.1016/S0140-6736(22)00089-7.

Peled, Y., Ram, E., Mandelboim, M., Lavee, J., Sternik, L., Segev, A., Wieder-Finesod, A., Halperin, R., Indenbaum, V., Levy, I., Patel, J., Raanani, E., Lustig, Y., & Rahav, G. (). Waning humoral immune response to the bnt162b2 vaccine in heart transplant recipients over 6 months. AMERICAN JOURNAL OF TRANSPLANTATION,. doi:10.1111/ajt.16998.

Tartof, S. Y., Slezak, J. M., Fischer, H., Hong, V., Ackerson, B. K., Ranasinghe, O. N., Frankland, T. B., Ogun, O. A., Zamparo, J. M., Gray, S., Valluri, S. R., Pan, K., Angulo, F. J., Jodar, L., & McLaughlin, J. M. (2021). Effectiveness of mrna bnt162b2 covid-19 vaccine up to 6 months in a large integrated health system in the usa: a retrospective cohort study. LANCET, 398, 1407–1416. doi:10.1016/S0140-6736(21)02183-8.

